# Mapping U.S. POINTER Cognitive-Slope Gains Onto Predicted Clinical Progression: An External-Cohort Translation Analysis With Exploratory Economic Thresholds

**DOI:** 10.64898/2026.05.17.26353395

**Authors:** Saki Nakashima, Kenichiro Sato, Yoshiki Niimi, Wataru Satake, Takeshi Iwatsubo, Alzheimer’s Disease Neuroimaging Initiative

## Abstract

**Background:** U.S. POINTER reported a modest structured-versus-self-guided difference in the annual rate of global cognitive change. However, the clinical and economic implications of this incremental standardized cognitive-slope benefit for delaying progression from cognitively normal status to mild cognitive impairment or dementia remain uncertain.

**Objectives:** To translate the U.S. POINTER cognitive-slope benefit into clinically interpretable progression outcomes in ADNI, A4, and LEARN, and to summarize scenario-based economic implications in ADNI subgroups.

**Design:** External-cohort translation analysis using two complementary analytic frameworks: an early-change landmark Cox model targeting Month 24 with a prespecified fallback window and a joint longitudinal–survival model.

**Setting:** ADNI, A4, and LEARN.

**Participants:** Cognitively normal participants. Landmark analytic samples included 399 ADNI participants with 61 events, 124 A4 participants with 37 events, and 394 LEARN participants with 45 events. Joint-model samples included 486 ADNI participants with 86 events, 1,064 A4 participants with 410 events, and 505 LEARN participants with 87 events.

**Intervention:** No multidomain lifestyle intervention was administered in ADNI, A4, or LEARN. ADNI and LEARN were observational longitudinal cohorts, whereas A4 was a randomized solanezumab trial; the present analysis did not estimate solanezumab treatment effects. We evaluated a counterfactual +0.029 SD/year improvement in cohort-specific mPACC slope, corresponding to the structured-versus-self-guided cognitive-slope difference reported in U.S. POINTER.

**Measurements:** The clinical outcome was sustained progression to mild cognitive impairment or dementia. Main translated measures were hazard ratios (HRs), 5-year risk differences (RDs), number needed to treat (NNT), and restricted mean survival time (RMST) differences. ADNI subgroup economic summaries used incremental 2-year delivery-cost scenarios and prespecified willingness-to-pay thresholds for prevented progression events and MCI-free years.

**Results:** Landmark analyses yielded small translated effects. For the +0.029 SD/year slope shift, HRs were 0.972 (95% CI, 0.949–0.989) in ADNI, 0.998 (0.989–1.005) in A4, and 0.996 (0.990–1.003) in LEARN, with corresponding 5-year RDs of 0.31 percentage points (95% CI, 0.12–0.57), 0.06 (-0.13 to 0.27), and 0.08 (-0.05 to 0.20). Joint models produced larger effects, with HRs of 0.831 (95% CrI, 0.776–0.879), 0.917 (0.907–0.927), and 0.833 (0.746–0.907), and 5-year RDs of 1.26 percentage points (0.90–1.68), 3.04 (2.65–3.43), and 2.25 (1.24–3.45), respectively. Corresponding NNT values were 79.1, 32.9, and 44.5, and RMST gains were 0.297, 1.242, and 0.617 months. In exploratory ADNI subgroup analyses, the small joint-model *APOE*-ε4+ & Aβ+ subgroup (61 participants, 22 events) showed the largest translated clinical effect, with HR 0.775 (95% CrI, 0.597–0.919), RD 2.65 percentage points (0.93–4.82), NNT 37.7, and RMST gain 0.723 months. In an exploratory threshold exercise, assuming an incremental 2-year delivery cost of $400 per participant for a structured intervention relative to a self-guided/reference intervention and a willingness-to-pay threshold of $100,000 per prevented progression event, the largest threshold-based net monetary benefit was observed in the *APOE*-ε4+ & Aβ+ joint-model subgroup (+$2,250/person). On an MCI-free-year basis under the same incremental-cost assumption, this subgroup also had the largest threshold-based net monetary benefit (+$5,622/person). These values should be interpreted as scenario-dependent thresholds rather than empirical cost-effectiveness estimates.

**Conclusions:** A U.S. POINTER–referenced structured-versus-self-guided cognitive-slope increment translated into directionally consistent reductions in predicted progression risk across ADNI, A4, and LEARN. The absolute clinical delay was generally modest and varied with cohort risk structure, biomarker/genotype enrichment, and analytic framework. Exploratory economic-threshold results suggested more favorable margins in higher-risk ADNI subgroups under low incremental-cost and high willingness-to-pay assumptions, but these findings should be interpreted as hypothesis-generating translation estimates rather than empirical cost-effectiveness evidence.

## 1. Introduction

Multidomain lifestyle interventions have emerged as an important strategy for dementia prevention because they can simultaneously address multiple modifiable risk factors (1–4). In the FINGER trial, a multidomain intervention incorporating diet, physical exercise, cognitive training, and vascular-risk monitoring slowed cognitive decline in at-risk older adults (2,5). Building on this framework, the U.S. Study to Protect Brain Health Through Lifestyle Intervention to Reduce Risk (POINTER) evaluated a similar approach in a large U.S. community-based population and showed that a structured multidomain intervention produced a statistically significant improvement in the annual rate of global cognitive change compared with a self-guided program (1,4). A major challenge, however, is that differences in the trajectory of a cognitive composite are not readily interpretable for clinicians, patients, or policymakers (6). In the U.S. POINTER report, the structured intervention showed a +0.029 SD/year between-group benefit in the annual rate of change of the trial’s primary global cognitive composite (1). Although statistically informative, this standardized slope difference does not directly indicate how much longer an individual might remain cognitively normal before progressing to mild cognitive impairment or dementia (6). In the external cohorts used here, the longitudinal cognitive outcome was modified Preclinical Alzheimer Cognitive Composite (mPACC), a composite intended to capture subtle early cognitive change across domains (7,8). A related translational challenge has been addressed in recent observational work linking modifiable behaviors to clinically interpretable cognitive and functional timelines. For example, an earlier literature used HABS data to relate physical activity to slower amyloid/tau accumulation and delayed cognitive and functional decline, illustrating how risk-factor associations can be expressed in time-based terms rather than only in standardized cognitive units (9). However, the clinical translation of an RCT-reported cognitive-slope difference, such as the incremental structured-versus-self-guided effect in U.S. POINTER, remains less well characterized.

To place this incremental structured-versus-self-guided cognitive effect into a clinically interpretable context, we used three external longitudinal cohorts: Alzheimer’s Disease Neuroimaging Initiative (ADNI), Anti-Amyloid Treatment in Asymptomatic Alzheimer’s Disease (A4), and Longitudinal Evaluation of Amyloid Risk and Neurodegeneration (LEARN), which represent complementary biomarker-risk settings (7,10–12). These cohorts were not selected to reproduce the U.S. POINTER target population. Rather, they were used to examine how the same standardized cognitive-slope increment maps onto progression metrics under different baseline risk, biomarker-enrichment, and follow-up structures.

We also considered the potential economic implications of such clinical translation as an exploratory threshold exercise. This was not intended to estimate the cost-effectiveness of lifestyle intervention itself. Instead, it asked what incremental delivery cost for a structured program, relative to a self-guided/reference program, could be justified under explicit willingness-to-pay assumptions (13,14). Accordingly, this study aimed to translate the POINTER-referenced structured-versus-self-guided cognitive-slope difference into predicted clinical progression metrics in ADNI, A4, and LEARN using two complementary statistical approaches, and to explore threshold-based economic margins in ADNI subgroups defined by *APOE* and amyloid status.

## 2. Methods

### 2.1 Study design and data sources

This study was designed as an external-cohort translation analysis. We used the incremental cognitive benefit reported in U.S. POINTER, namely the +0.029 SD/year structured-versus-self-guided between-group difference in the annual rate of change of the trial’s primary global cognitive composite, as the reference standardized slope increment for external-cohort translation (1,4). Thus, the translated quantity represents the incremental structured-program advantage over a self-guided/reference program, not the total effect of lifestyle intervention versus no intervention.

The external cohorts were ADNI, A4, and LEARN (7,10–12). These cohorts were not intended to represent the U.S. POINTER target population directly, nor were they intended to replicate U.S. POINTER. Rather, they were selected because they provide longitudinal cognitive and clinical follow-up in cognitively normal older adults across distinct AD risk settings: ADNI provides an observational cohort with heterogeneous biomarker status, A4 provides an amyloid-enriched preclinical AD trial population, and LEARN provides a companion amyloid-non-elevated cohort. This design allowed us to examine how the same standardized cognitive-slope increment translated under different baseline risk and biomarker structures.

A4 differed from ADNI and LEARN because it was originally conducted as a randomized solanezumab trial. However, the present study did not evaluate solanezumab efficacy or use A4 as an intervention comparator. Instead, A4 and LEARN were used as longitudinal amyloid-elevated and amyloid-non-elevated progression datasets. This interpretation is consistent with the published A4 double-blind trial result, in which solanezumab did not slow cognitive decline compared with placebo over 240 weeks.

The longitudinal cognitive outcome was a cohort-specific mPACC or PACC-derived composite, standardized within cohort for analysis. In ADNI, mPACC was reconstructed from available cognitive tests using baseline cohort norms and expressed as change from each participant’s baseline composite score. In A4 and LEARN, we used the Alzheimer’s Clinical Trials Consortium (ACTC)-derived PACC variable, which was already expressed relative to each participant’s baseline PACC.raw value. Detailed component definitions and preprocessing rules are provided in Supplementary Methods.

Amyloid status was defined according to the original cohort-specific biomarker framework. In ADNI, amyloid positivity was operationalized using available CSF or amyloid PET information closest to baseline. In A4 and LEARN, amyloid status followed the screening florbetapir PET classification used for trial eligibility, with A4 representing an amyloid-elevated cohort and LEARN representing a companion amyloid-non-elevated cohort. Detailed assay- and tracer-specific cutoffs are provided in Supplementary Methods. Amyloid status was used for descriptive summaries and ADNI subgroup definitions, but not as a modeling variable in the main cross-cohort translation analyses.

The clinical outcome was sustained progression from cognitively normal status to mild cognitive impairment or dementia. Sustained conversion was defined as the first visit at which mild cognitive impairment or dementia was observed and confirmed again at the next visit. This rule was used to reduce the likelihood that a single fluctuating diagnosis would be counted as a true clinical event.

This translation requires an explicit causal interpretation assumption: the association between cognitive slope and subsequent clinical progression estimated in the external cohorts was assumed to approximate the progression effect that would be observed if an intervention produced a comparable improvement in cognitive slope. This assumption is not directly testable in the present data. Therefore, the analyses should be interpreted as counterfactual clinical translations of a slope shift, not as direct estimates of U.S. POINTER intervention effects in ADNI, A4, or LEARN.

### 2.2 Statistical models

We used two prespecified models to examine the same translation question under complementary levels of modeling complexity. The landmark Cox model provides a transparent two-stage summary based on early cognitive change (15). It first summarizes each participant’s cognitive slope during the early landmark window and then examines whether that slope predicts later clinical progression. This approach is straightforward to interpret and closely linked to an early-change estimand, but it discards longitudinal information after the landmark and treats the estimated early slope as if it were observed without error.

The joint longitudinal–survival model is more complex but better suited to repeated-measures data with noisy trajectories and irregular follow-up (16,17). It fits the cognitive trajectory and the time-to-event process simultaneously, uses all available repeated cognitive observations, and links the latent instantaneous rate of cognitive change to progression risk. We interpreted the landmark and joint-model analyses as complementary sensitivity frameworks for the same translation question, rather than as competing approaches intended to identify a single definitive estimate. Detailed model equations are provided in Supplementary Methods. A schematic overview of the landmark and joint longitudinal–survival modeling frameworks is provided in Figure S1.

The primary cohort-level translation focused on the direction and magnitude of the translated hazard ratio (HR) and 5-year risk difference (RD) under the +0.029 SD/year slope shift. Number needed to treat (NNT) and restricted mean survival time (RMST) were treated as clinically interpretable absolute summaries derived from the same fitted models. ADNI subgroup and economic analyses were prespecified as exploratory and were not used for confirmatory inference. Because the analyses were intended to quantify translation across cohorts and modeling frameworks rather than to test multiple independent hypotheses, we did not apply multiplicity-adjusted P values. Interval estimates should therefore be interpreted descriptively, particularly for subgroup analyses.

Because the published U.S. POINTER slope estimate was itself uncertain, we performed an approximate Monte Carlo sensitivity analysis propagating uncertainty in the reference effect. The slope increment was treated as Δ_POINTER ∼ Normal(0.029, SE_POINTER²), with SE_POINTER derived from the published 95% confidence interval. Because individual-level U.S. POINTER data were unavailable, this sensitivity analysis approximated uncertainty propagation using the primary estimates and intervals from Tables 2 and 3 rather than refitting the individual-level external-cohort models. Details are provided in Supplementary Methods.

### 2.3 Eligibility determination

For the landmark analysis, eligibility was defined at a Month 24 target landmark. Participants entered the landmark risk set only if they remained cognitively normal through that landmark; those who developed mild cognitive impairment or dementia before the landmark were excluded from the post-landmark analysis. The landmark visit was defined as the assessment closest to Month 24 within a prespecified window. We first searched for a visit between Month 24 and Month 36 and selected the one closest to Month 24. If no such visit was available, we used the nearest prior visit between Month 12 and less than Month 24. This fallback rule was intended to reduce unnecessary exclusion of participants who did not have an exact Month 24 assessment.

For the joint-model analysis, baseline was defined as the visit closest to Month 0 within a ±3-month window, and only participants who were cognitively normal at that baseline definition were included. Participants with missing baseline covariates required for model adjustment were excluded from the corresponding complete-case analytic sample. Missing longitudinal mPACC observations were not imputed; mixed-effects and joint-model analyses used available repeated measures under the corresponding model assumptions.

### 2.4 Landmark Cox analysis

For the landmark Cox analysis, cohort-specific mPACC was standardized to a z score, and each participant’s early annual cognitive slope was estimated from baseline through the landmark window. The estimated slope was then entered into a post-landmark Cox model adjusted for age, sex, education, and *APOE*-ε4. The POINTER-referenced translation was evaluated by comparing the observed-slope scenario with a shifted-slope scenario in which each participant’s slope was improved by +0.029 SD/year. From these scenarios, we derived the translated HR, 5-year RD, NNT, and 5-year RMST difference. Uncertainty was estimated using 500 participant-level bootstrap resamples that reran the full slope-estimation and Cox-model pipeline. Detailed equations and bootstrap procedures are provided in Supplementary Methods.

The proportional hazards assumption for the Cox survival components used in the landmark and joint-model analyses was assessed using Schoenfeld residuals. Results are summarized in Table S1.

### 2.5 Joint longitudinal–survival model

For the joint longitudinal–survival analysis, repeated mPACC observations from baseline through Month 84 were modeled jointly with time to sustained conversion. The longitudinal submodel included fixed effects for time, age, sex, education, and *APOE*-ε4, together with participant-specific random intercepts and slopes. The survival submodel included the same baseline covariates and linked the latent instantaneous slope of the cognitive trajectory to the event process. We used a slope-only association structure because the scientific question was how a difference in the rate of cognitive change would translate into a difference in conversion risk.

Models were estimated in a Bayesian framework using *JMbayes2*. Posterior predictive survival trajectories were used to estimate the translated HR, RD, NNT, and RMST under observed-slope and shifted-slope scenarios. Detailed equations, Markov chain Monte Carlo settings, convergence diagnostics, and posterior prediction procedures are provided in Supplementary Methods.

### 2.6 ADNI sensitivity analysis aligned to public U.S. POINTER parent-trial marginals

Because the primary analysis examined how a U.S. POINTER–sized improvement in cognitive slope would translate within the observed covariate distributions of ADNI, A4, and LEARN, rather than by reconstructing POINTER-like individuals within those cohorts, we conducted an ADNI-only sensitivity analysis in which selected baseline marginal distributions were aligned with the published U.S. POINTER parent-trial distribution using entropy-balancing weights (18,19). Calibration targets included age, sex, age ≥70 years, and *APOE*-ε4 carrier status; amyloid positivity was not included because the published value corresponded to an imaging ancillary cohort rather than the full randomized parent trial.

For landmark analyses, weights were applied in the Cox-model translation. For joint models, a weighted pseudo-population was generated because the JMbayes2 implementation did not directly accommodate participant-level calibration weights. This analysis was interpreted as marginal realignment, not reconstruction of individual-level U.S. POINTER participants. Reweighting diagnostics are provided in Table S2.

### 2.7 ADNI subgroup economic analysis

Economic analyses were restricted to ADNI because A4 and LEARN were largely defined by amyloid-screening eligibility and were therefore not suitable for balanced biomarker-based subgroup comparisons. We evaluated ADNI overall, *APOE*-ε4 carriers, amyloid-positive participants, and *APOE*-ε4 plus amyloid-positive participants.

The economic component was designed as an exploratory cost-threshold analysis rather than a within-trial cost-effectiveness analysis (20,21). We did not estimate quality-adjusted life-years, observed implementation costs, health-care utilization, or societal costs from U.S. POINTER. Instead, we estimated the maximum incremental 2-year delivery cost of a structured program, relative to a self-guided/reference program, that would be consistent with prespecified willingness-to-pay assumptions.

The base incremental-cost scenario was $400 per participant over 2 years, with $600 and $800 examined in sensitivity analyses. These amounts were intended to represent illustrative incremental delivery-cost margins for the additional structured-program intensity and not the full cost of delivering a multidomain lifestyle intervention. WTP thresholds were used only as illustrative scale-setting assumptions and are not dimensionally equivalent to conventional cost-per-QALY thresholds. Economic formulas and scenario definitions are provided in Table S3 and Supplementary Methods.

### 2.8 Ethics

This study was approved by the University of Tokyo Graduate School of Medicine institutional ethics committee (ID: 2025264NI). Informed consent was not required because this study used de-identified secondary data obtained through established data-sharing repositories. This report was prepared with reference to the STROBE reporting recommendations where applicable.

## 3. Results

### 3.1 Basic Characteristics

The baseline CN samples comprised 540 participants with 86 sustained progression events in ADNI, 1,169 participants with 410 events in A4, and 538 participants with 87 events in LEARN. Mean age in these baseline CN samples was 73.3 years in ADNI, 71.9 years in A4, and 70.5 years in LEARN; female proportions were 52.8%, 40.6%, and 38.7%, respectively; mean years of education were 16.4, 16.6, and 16.8; and mean MMSE values were 29.1, 28.8, and 29.0. The landmark analytic samples included 399 participants with 61 events in ADNI, 124 participants with 37 events in A4, and 394 participants with 45 events in LEARN. The corresponding joint-model analytic datasets included 486 participants with 86 events in ADNI, 1,064 participants with 410 events in A4, and 505 participants with 87 events in LEARN (Table 1).

**Table 1.** Baseline characteristics of the analytic samples across cohorts.

| Cohort | Sample | N | Sustained<br>MCI/dementia<br>events, n | Age,<br>mean<br>(SD) | Female, % | Education<br>years,<br>mean (SD) | MMSE,<br>mean<br>(SD) | APOE-ε4<br>carrier, % | Amyloid<br>positive, % |
| --- | --- | --- | --- | --- | --- | --- | --- | --- | --- |
| ADNI | Baseline CN | 540 | 86 | 73.3<br>(6.4) | 52.8% | 16.4 (2.6) | 29.1 (1.1) | 27.7% | 42.9% |
|  | Landmark<br>model | 399 | 61 | 73.9<br>(6.1) | 50.9% | 16.4 (2.6) | 29.2 (1.1) | 27.0% | 44.3% |
|  | Joint model | 486 | 86 | 73.8<br>(6.1) | 51.9% | 16.4 (2.7) | 29.1 (1.1) | 27.5% | 44.8% |
| A4 | Baseline CN | 1169 | 410 | 71.9<br>(4.8) | 40.6% | 16.6 (2.8) | 28.8 (1.3) | 58.9% | 94.8% |
|  | Landmark<br>model | 124 | 37 | 71.7<br>(4.5) | 37.9% | 16.9 (2.5) | 29.0 (1.1) | 51.6% | 92.7% |
|  | Joint model | 1064 | 410 | 71.8<br>(4.8) | 41.0% | 16.6 (2.8) | 28.8 (1.3) | 59.5% | 95.1% |
| LEARN | Baseline CN | 538 | 87 | 70.5<br>(4.3) | 38.7% | 16.8 (2.6) | 29.0 (1.2) | 22.9% | 0.7% |
|  | Landmark<br>model | 394 | 45 | 70.0<br>(4.0) | 35.8% | 16.8 (2.6) | 29.0 (1.2) | 22.4% | 0.5% |
|  | Joint model | 505 | 87 | 70.5<br>(4.3) | 39.0% | 16.8 (2.6) | 29.0 (1.2) | 22.3% | 0.8% |

Abbreviations: ADNI, Alzheimer's Disease Neuroimaging Initiative; A4, Anti-Amyloid
Treatment in Asymptomatic Alzheimer's Disease; LEARN, Longitudinal Evaluation of
Amyloid Risk and Neurodegeneration; MMSE, Mini-Mental State Examination.
MCI/dementia events indicate the number of participants with sustained progression to MCI or
dementia during follow-up within each analytic sample. For the landmark sample, events are
counted after landmark entry.

Amyloid positivity differed markedly by cohort and was stable across analytic datasets. In the joint-model sample, amyloid positivity was 44.8% in ADNI, 95.1% in A4, and 0.8% in LEARN. The same pattern was present in the baseline CN and landmark samples, indicating that A4 and LEARN were informative for contrasting biomarker settings but were not suitable for balanced biomarker-based subgroup comparisons.

The A4 landmark sample was substantially smaller than the baseline CN sample because few participants had eligible observations near the prespecified 24-month landmark. In A4, only 147 participants had an assessment within the 12- to 36-month landmark window, of whom 124 remained cognitively normal and were included in the final landmark analysis. Among these 124 participants, 123 (99.2%) entered through the nearest prior visit within Month 12 to <24 rather than through a Month 24–36 visit. Therefore, A4 landmark estimates should be interpreted as early-window landmark estimates rather than as estimates based on a true Month 24 landmark assessment. The distribution of landmark-window selection modes across cohorts is shown in Table S4.

### 3.2 Primary cohort-level translation results

In the landmark analysis, translated absolute effects were small across cohorts. Standardized to the +0.029 SD/year slope improvement, HRs were 0.972 (95% CI 0.949-0.989) in ADNI, 0.998 (0.989-1.005) in A4, and 0.996 (0.990-1.003) in LEARN (Table 2). The corresponding 5-year RDs were 0.31 (95% CI 0.12-0.57), 0.06 (-0.13 to 0.27), and 0.08 (-0.05 to 0.20) percentage points, respectively.

**Table 2.** Cohort-level translated clinical effects standardized to a +0.029 SD/year improvement in cognitive slope.

| Cohort | Model | HR (95% interval) | RD (95% interval, pp) | NNT (95% interval) | RMST (95% interval, month) |
| --- | --- | --- | --- | --- | --- |
| ADNI | Landmark model | 0.972 (0.949-0.989) | 0.31 (0.12-0.57) | 322.9<br>(178.1-1140.8) | 0.062 (0.016-0.114) |
|  | Joint model | 0.831 (0.776-0.879) | 1.26 (0.90-1.68) | 79.1 (59.5-111.2) | 0.297 (0.211-0.394) |
| A4 | Landmark model | 0.998 (0.989-1.005) | 0.06 (-0.13-0.27) | Unstable | 0.013<br>(-0.032-0.065) |
|  | Joint model | 0.917 (0.907-0.927) | 3.04 (2.65-3.43) | 32.9 (29.2-37.7) | 1.242 (1.086-1.398) |
| LEARN | Landmark model | 0.996 (0.990-1.003) | 0.08 (-0.05-0.20) | Unstable | 0.020<br>(-0.013-0.052) |
|  | Joint model | 0.833 (0.746-0.907) | 2.25 (1.24-3.45) | 44.5 (29.0-80.6) | 0.617 (0.341-0.944) |

Abbreviations: HR, hazard ratio; RD, risk difference; NNT, number needed to treat; RMST, restricted mean survival time; CI, confidence interval; CrI, credible interval. Intervals are 95% confidence intervals for landmark models and 95% credible intervals for joint models.

In the joint-model analysis, the translated relative and absolute effects were larger than in the landmark analysis, although absolute RMST gains remained modest. For the +0.029 SD/year shift, HRs were 0.831 (95% CrI 0.776-0.879) in ADNI, 0.917 (0.907-0.927) in A4, and 0.833 (0.746-0.907) in LEARN. The corresponding 5-year RDs were 1.26 (95% CrI 0.90-1.68), 3.04 (2.65-3.43), and 2.25 (1.24-3.45) percentage points, with NNT values of 79.1, 32.9, and 44.5 and RMST gains of 0.297, 1.242, and 0.617 months, respectively (Table 2).

Joint-model convergence diagnostics did not indicate major convergence problems in the final fitted models. For each cohort, the maximum R-hat across monitored parameters was below the prespecified threshold of 1.10. Effective sample sizes and model-specific time-function choices are summarized descriptively in Table S5. No model required interpretation based on a non-converged primary fit.

### 3.3 ADNI-only sensitivity analysis aligned to published U.S. POINTER parent-trial marginals

Reweighting ADNI toward the published U.S. POINTER parent-trial marginals moved the observed baseline CN distribution in the expected direction: mean age from 73.7 to 68.7 years, female proportion from 52.3% to 67.7%, age 70 years or older from 75.2% to 44.7%, and *APOE*-ε4 positivity from 27.7% to 29.8%; the effective sample size was 254.6 (Table S2).

This baseline realignment attenuated but did not reverse the translated benefit. In the landmark framework, the 5-year RD decreased from 0.31 to 0.21 percentage points, the NNT increased from 322.9 to 482.2, and the RMST gain decreased from 0.062 to 0.039 months. In the joint-model framework, the HR changed from 0.831 to 0.877, the 5-year RD from 1.26 to 0.86 percentage points, the NNT from 79.1 to 116.0, and the RMST gain from 0.297 to 0.241 months (Table 4).

**Table 3.**
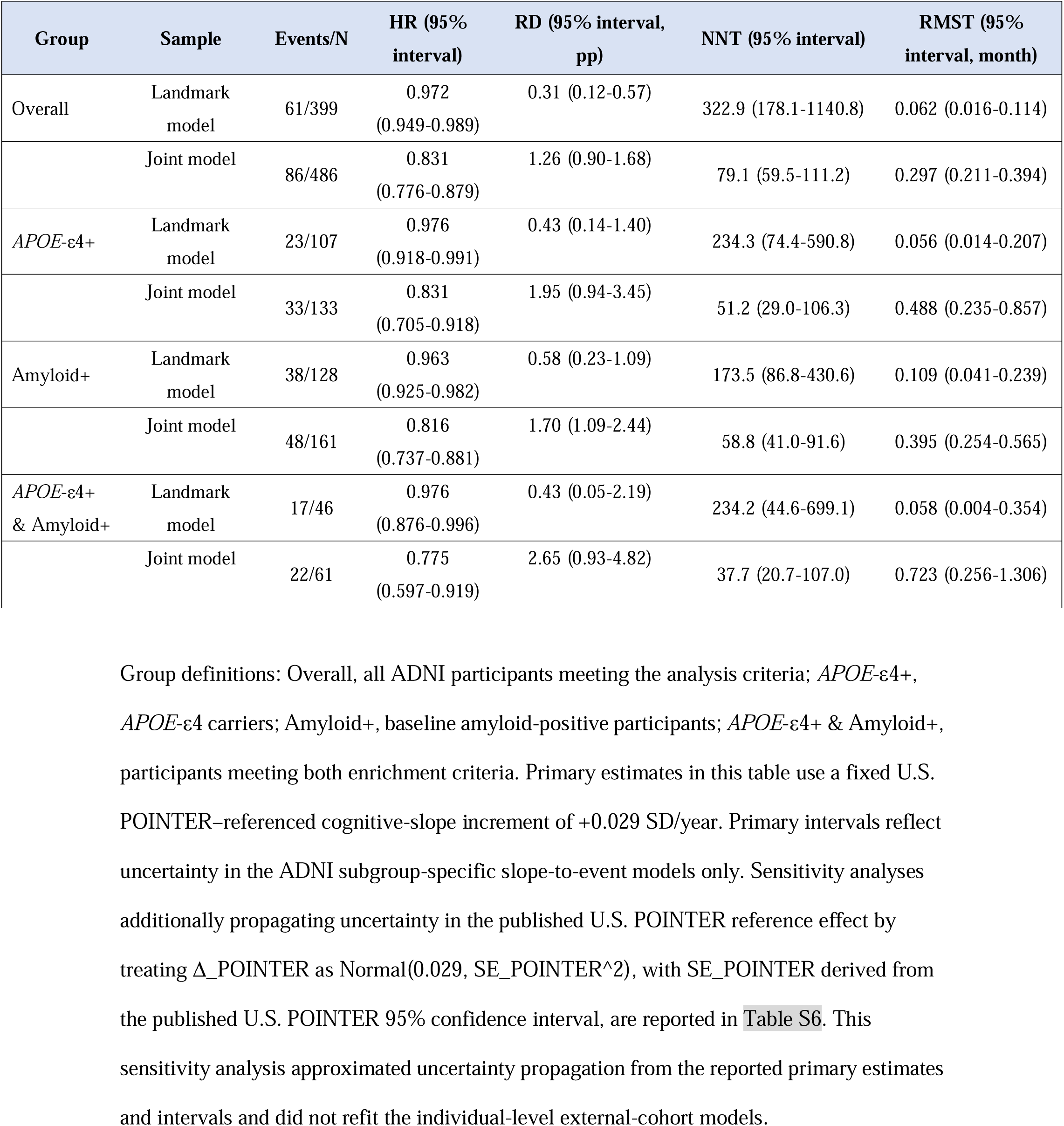
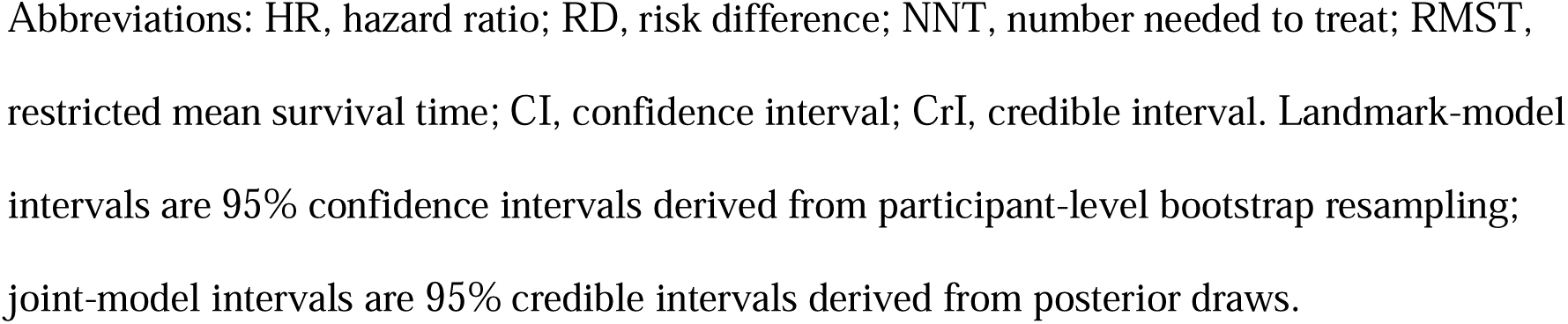
ADNI subgroup translated clinical effects.

| Group | Sample | Events/N | HR (95% interval) | RD (95% interval, pp) | NNT (95% interval) | RMST (95% interval, month) |
| --- | --- | --- | --- | --- | --- | --- |
| Overall | Landmark model | 61/399 | 0.972<br>(0.949-0.989) | 0.31 (0.12-0.57) | 322.9 (178.1-1140.8) | 0.062 (0.016-0.114) |
|  | Joint model | 86/486 | 0.831<br>(0.776-0.879) | 1.26 (0.90-1.68) | 79.1 (59.5-111.2) | 0.297 (0.211-0.394) |
| *APOE*-ε4+ | Landmark model | 23/107 | 0.976<br>(0.918-0.991) | 0.43 (0.14-1.40) | 234.3 (74.4-590.8) | 0.056 (0.014-0.207) |
|  | Joint model | 33/133 | 0.831<br>(0.705-0.918) | 1.95 (0.94-3.45) | 51.2 (29.0-106.3) | 0.488 (0.235-0.857) |
| Amyloid+ | Landmark model | 38/128 | 0.963<br>(0.925-0.982) | 0.58 (0.23-1.09) | 173.5 (86.8-430.6) | 0.109 (0.041-0.239) |
|  | Joint model | 48/161 | 0.816<br>(0.737-0.881) | 1.70 (1.09-2.44) | 58.8 (41.0-91.6) | 0.395 (0.254-0.565) |
| *APOE*-ε4+ & Amyloid+ | Landmark model | 17/46 | 0.976<br>(0.876-0.996) | 0.43 (0.05-2.19) | 234.2 (44.6-699.1) | 0.058 (0.004-0.354) |
|  | Joint model | 22/61 | 0.775<br>(0.597-0.919) | 2.65 (0.93-4.82) | 37.7 (20.7-107.0) | 0.723 (0.256-1.306) |

Abbreviations: HR, hazard ratio; RD, risk difference; NNT, number needed to treat; RMST, restricted mean survival time; CI, confidence interval; CrI, credible interval. Landmark-model intervals are 95% confidence intervals derived from participant-level bootstrap resampling; joint-model intervals are 95% credible intervals derived from posterior draws.

**Table 4.** ADNI-only sensitivity analysis reweighted to the published U.S. POINTER parent-trial baseline marginals.

| Analysis | HR (95% interval) | 5-year RD (95% interval, pp) | NNT (95% interval) | RMST difference (95% interval, month) |
| --- | --- | --- | --- | --- |
| Main landmark model | 0.972 (0.949-0.989) | 0.31 (0.12-0.57) | 322.9 (178.1-1140.8) | 0.062 (0.016-0.114) |
| Rewighted landmark model | 0.972 (0.948-0.991) | 0.21 (0.06-0.44) | 482.2 (226.9-1704.9) | 0.039 (0.009-0.096) |
| Main joint model | 0.831 (0.776-0.879) | 1.26 (0.90-1.68) | 79.1 (59.5-111.2) | 0.297 (0.211-0.394) |
| Rewighted joint model | 0.877 (0.833-0.915) | 0.86 (0.59-1.17) | 116.0 (85.6-168.3) | 0.241 (0.166-0.326) |

The reweighted analyses align ADNI baseline marginals for age, sex, age $\geq 70$ years, and *APOE-ε4* positivity to the published U.S. POINTER parent-trial distribution. They do not reconstruct individual-level POINTER participants and should be interpreted as a sensitivity analysis based on marginal realignment only. For landmark analyses, intervals are 95% confidence intervals derived from participant-level bootstrap resampling. For joint-model analyses, intervals are 95% credible intervals derived from posterior draws.
Abbreviations: HR, hazard ratio; RD, risk difference; NNT, number needed to treat; RMST, restricted mean survival time.

### 3.4 ADNI subgroup translation results

Within ADNI, subgroup enrichment changed the size of the translated benefit. In the landmark analysis, subgroup-specific HRs ranged from 0.963 to 0.976, and 5-year risk differences ranged from 0.31 to 0.58 percentage points. NNT values were 322.9 in the overall sample, 234.3 in *APOE*-ε4+, 173.5 in amyloid+, and 234.2 in *APOE*-ε4+ plus amyloid+ participants. The corresponding RMST gains were 0.062, 0.056, 0.109, and 0.058 months, respectively. Landmark 95% interval estimates are shown in Table 3.

The joint-model subgroup analysis produced larger translated effects than the landmark analysis, but the absolute RMST gains remained below 1 month across ADNI subgroups. HRs were 0.831 (95% CrI 0.705-0.918) in *APOE*-ε4+, 0.816 (0.737-0.881) in amyloid+, and 0.775 (0.597-0.919) in *APOE*-ε4+ plus amyloid+ participants, compared with 0.831 (0.776-0.879) in the overall sample. The corresponding 5-year risk differences were 1.26, 1.95, 1.70, and 2.65 percentage points in the overall, *APOE*-ε4+, amyloid+, and *APOE*-ε4+ plus amyloid+ groups, respectively. NNT improved to 79.1 in the overall sample, 51.2 in *APOE*-ε4+, 58.8 in amyloid+, and 37.7 in *APOE*-ε4+ plus amyloid+ participants. The corresponding RMST gains were 0.297, 0.488, 0.395, and 0.723 months. Across exploratory ADNI subgroups, the largest translated effects were observed in the *APOE*-ε4+ plus amyloid+ subgroup (Table 3 and Figure 1A). However, this subgroup was small, particularly in the landmark analysis, and interval estimates were wide. Therefore, these subgroup findings should be interpreted as enrichment-related signals from a scenario-based translation analysis, not as evidence for preferential implementation in this subgroup.

**Figure 1.**
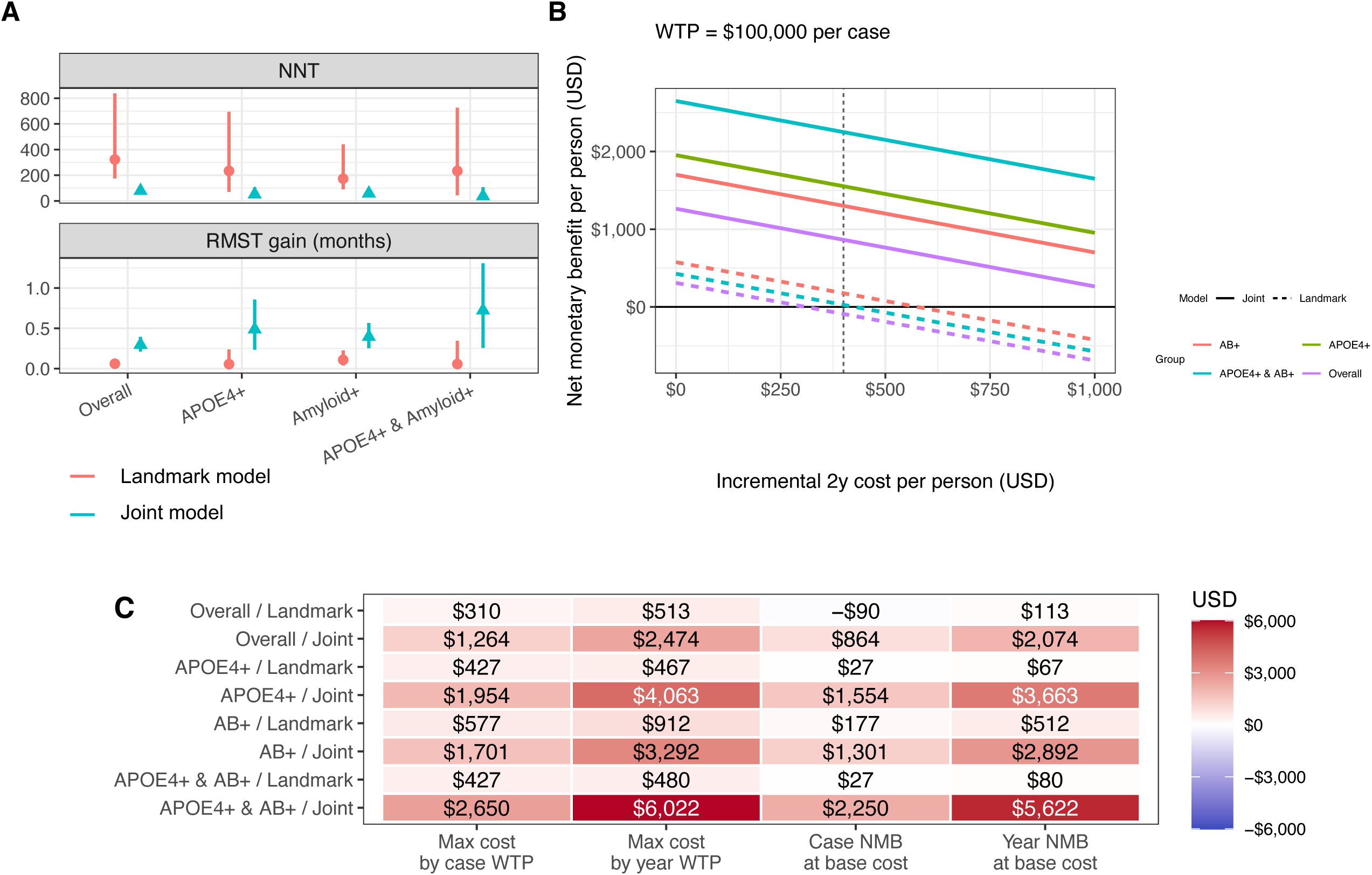
ADNI subgroup translated clinical and economic effects under a U.S. POINTER–referenced cognitive-slope shift. This figure summarizes the clinical and economic implications of applying a counterfactual +0.029 SD/year improvement in mPACC slope to ADNI subgroups. This slope shift corresponds to the structured-versus-self-guided cognitive slope difference reported in U.S. POINTER. Analyses were performed in ADNI overall and in three enriched subgroups: *APOE*-ε4+, amyloid-positive (Aβ+), and *APOE*-ε4+ & Aβ+. In panel A, red circles indicate the landmark Cox model and blue triangles indicate the joint longitudinal–survival model. In panel B, color indicates subgroup and line type indicates model. In panel C, rows indicate subgroup-model combinations. A, Translated clinical effects. The upper panel shows the 5-year number needed to treat (NNT) to prevent one conversion from cognitively normal status to MCI or dementia. The lower panel shows the 5-year restricted mean survival time (RMST) gain, expressed in months of additional MCI-free survival. Estimates are shown for each ADNI subgroup under the +0.029 SD/year mPACC slope shift. Vertical lines indicate uncertainty intervals. Lower NNT and higher RMST gain indicate larger translated clinical benefit. B, Case-prevention threshold plot. The x-axis shows the assumed incremental 2-year delivery cost per person in USD for the structured intervention relative to a self-guided/reference intervention. The y-axis shows case-based net monetary benefit (NMB) per person, calculated as: *NMB = (WTP per prevented case / NNT) − incremental cost*. The willingness-to-pay threshold was set at $100,000 per prevented case. The vertical dashed line indicates the base incremental cost assumption of $400 per person over 2 years. Values above zero indicate that the translated clinical benefit exceeds the assumed intervention cost on a case-prevention basis. C, ADNI subgroup economic summary at the base incremental cost. The heatmap summarizes per-person threshold-based economic values in USD at the base incremental 2-year delivery cost of $400 per person. The case-based columns use a WTP threshold of $100,000 per prevented case, and the MCI-free-year columns use a WTP threshold of $100,000 per MCI-free year. “Max cost by case WTP” is the maximum 2-year cost per person that would be justified at $100,000 per prevented case, calculated as WTP/NNT. “Max cost by year WTP” is the maximum 2-year cost per person that would be justified at $100,000 per MCI-free year, calculated from the RMST gain. “Case NMB at base cost” and “Year NMB at base cost” subtract the base cost of $400 from these two maximum-cost values, respectively. Red indicates positive values, blue indicates negative values, and white indicates zero. Positive NMB indicates that the translated benefit exceeds the base cost under the specified willingness-to-pay assumption.

### 3.5 ADNI subgroup economic translation

Figure 1B summarizes the case-based threshold analysis, in which net monetary benefit per person is plotted against incremental 2-year cost per person using a willingness-to-pay threshold of 100,000 USD per case prevented. On this basis, the break-even incremental cost was lowest in the overall landmark model and highest in the *APOE*-ε4+ plus amyloid+ joint model, with the remaining subgroup-model combinations ordered between these extremes.

At the illustrative base scenario of 400 USD incremental cost per participant over 2 years, the largest positive case-based threshold margin was observed in the *APOE*-ε4+ plus amyloid+ joint-model subgroup (+2,250 USD/person). Other joint-model subgroup margins were also positive: +1,554 USD/person in *APOE*-ε4+, +1,301 USD/person in amyloid+, and +864 USD/person in the overall ADNI sample. In contrast, landmark-model margins were small and ranged from -90 USD/person in the overall sample to +177 USD/person in the amyloid+ subgroup. These values reflect threshold-based calculations from modeled risk differences, not observed economic outcomes.

On an MCI-free-year basis using the illustrative threshold of 100,000 USD per MCI-free year, all subgroup-model combinations remained above break-even at the 400 USD incremental-cost scenario. However, these results were mechanically driven by the assumed WTP threshold, the modeled RMST gain, and the assumed incremental cost. Alternative fixed incremental cost scenarios of 600 and 800 USD per person over 2 years, combined with case-based WTP thresholds of 100,000 and 150,000 USD per case prevented, are summarized in Figure S2.

## 4. Discussion

This study translated the U.S. POINTER structured-versus-self-guided cognitive-slope difference into predicted clinical progression metrics in ADNI, A4, and LEARN (1,22,23). The translated quantity is the incremental advantage of a more structured program over a self-guided/reference program, not the total effect of lifestyle intervention versus no intervention. Across cohorts, a +0.029 SD/year slope increment translated into directionally consistent reductions in predicted progression risk, with absolute effects that varied by baseline risk, biomarker structure, follow-up design, and modeling framework (10–12,24).

The contrast between the landmark and joint-model analyses is clinically informative, but the two frameworks should not be interpreted as estimating identical quantities (15–17). The landmark Cox model used a 24-month risk-set definition, excluded individuals who progressed before the landmark, and related an estimated early cognitive slope to subsequent post-landmark progression. In contrast, the joint longitudinal–survival model started at baseline, used all available repeated cognitive observations during follow-up, and linked the latent instantaneous slope of the cognitive trajectory to the event process. Therefore, differences between the two models reflect not only measurement-error handling, but also differences in risk-set construction, information use, and model assumptions.

The larger translated effects in the joint models are consistent with two nonexclusive explanations. First, the landmark model uses an estimated early observed slope as a second-stage predictor; measurement error in this estimated slope can attenuate the slope-to-event association and move the translated HR toward the null. Second, the joint model links a latent instantaneous slope to the event process and can therefore be more sensitive to slope-related risk, but this sensitivity depends on the slope-only association structure and other modeling assumptions. The divergence was especially important in A4, where nearly all landmark participants entered through the prespecified 12- to <24-month fallback window rather than a true Month 24–36 visit. We therefore present the two frameworks as complementary sensitivity analyses rather than as competing estimates of a single definitive effect. The divergence between the two approaches is itself an important result, because it shows that the translated clinical benefit of a POINTER-sized cognitive-slope shift depends materially on risk-set construction, measurement-error handling, and model structure.

Viewed on an absolute time scale, even the joint-model estimates imply modest individual-level delay over 5 years. The translated RMST gains were approximately 0.3 months in ADNI, 1.2 months in A4, and 0.6 months in LEARN. Thus, the main implication is not that a POINTER-sized cognitive-slope shift would produce a large short-term delay in clinical progression for each individual, but that standardized cognitive-slope effects can be translated into progression metrics that quantify the expected scale of benefit. This distinction is important for interpreting prevention trials, where small individual-level effects may still be relevant if an intervention is low cost, scalable, safe, and targeted to populations with sufficiently high baseline risk.

The ADNI reweighting analysis provides a further caution for interpretation. When ADNI was aligned to selected published U.S. POINTER parent-trial marginals, the joint-model translation was attenuated, with the HR moving from 0.831 to 0.877 and the 5-year RD decreasing from 1.26 to 0.86 percentage points. This suggests that the slope-to-progression translation estimated in the older, more ADNI-like distribution may not directly apply to a younger and more female POINTER-like marginal distribution. Because the reweighting used only selected published marginals and could not reconstruct the joint distribution of individual-level POINTER participants, this analysis should be interpreted as directional evidence of sensitivity to target-population structure rather than as formal transportability.

This interpretation is consistent with the broader multidomain dementia-prevention literature, in which cognitive effects have generally been modest and have varied according to population selection, intervention intensity, delivery mode, and baseline risk (2,25,26). FINGER showed cognitive benefit in at-risk older adults (2), whereas MAPT and preDIVA reported more limited or context-dependent effects in broader or differently selected populations (25,26). More recent multidomain prevention work, including the online Maintain Your Brain trial and pooled MAPT/preDIVA responder analyses (27,28), further emphasizes that intervention effects and responder profiles depend strongly on implementation context and participant selection. In parallel, recent observational work has begun translating modifiable risk-factor associations into clinically interpretable time-based outcomes, such as delayed cognitive and functional decline in preclinical AD. The present analysis differs by mapping an RCT-reported structured-versus-self-guided cognitive-slope difference onto progression metrics in external cohorts, comparing two modeling frameworks, and adding an exploratory economic-threshold layer.

Biomarker structure is a particularly important source of this heterogeneity (10,11,24). A4 was designed to enroll cognitively normal individuals with elevated amyloid on screening PET, whereas LEARN was designed as the companion cohort of otherwise similar individuals without elevated amyloid (10,11). These design features make A4 and LEARN highly informative for examining how a common cognitive-slope shift behaves in contrasting biomarker settings, but they also make them unsuitable for balanced subgroup-based economic comparison. For this reason, we restricted subgroup economic translation to ADNI, where amyloid positive and amyloid-negative participants, as well as *APOE*-ε4-enriched and non-enriched participants, were all represented within the same observational framework.

The economic analysis should be interpreted as an exploratory threshold exercise rather than as direct within-trial cost-effectiveness evidence (22,23,29,30). We did not estimate quality-adjusted life-years, observed implementation costs, health-care utilization, or societal costs from POINTER. Instead, we asked what incremental delivery cost would be justified if the modeled progression benefit were valued at prespecified WTP thresholds. Under these assumptions, the *APOE*-ε4+ plus amyloid-positive ADNI subgroup had the largest positive threshold margin, especially in the joint-model framework. However, this result should be interpreted as identifying where a favorable economic profile might arise under assumed costs and WTP thresholds, not as evidence that intensive lifestyle delivery is cost-effective in that subgroup or that the subgroup should be preferentially targeted for implementation.

This threshold framing differs from formal QALY-based cost-effectiveness analyses of multidomain prevention programs. For example, prior economic evaluations of FINGER (31), Maintain Your Brain (32), and J-MINT (33) used observed or modeled costs and QALY-related outcomes to estimate cost-effectiveness from health-care or societal perspectives. In contrast, the present study did not observe program costs, health-care utilization, utilities, or QALYs. Moreover, the WTP thresholds used for prevented progression events and MCI-free years are not dimensionally equivalent to conventional cost-per-QALY thresholds and should be interpreted only as illustrative decision thresholds. Its economic component should therefore be read as a decision-threshold exercise asking how large the incremental structured-program cost could be under assumed WTP values, conditional on the translated clinical effect. This distinction is important because the assumed $400 base scenario represents an illustrative low incremental delivery-cost margin relative to a self-guided/reference program and not the full cost of implementing a multidomain lifestyle intervention.

Recent A4/LEARN Medicare-linked evidence also supports the relevance of clinical progression, rather than amyloid elevation alone, for economic interpretation (34). In that analysis, cognitively unimpaired A4 Medicare participants as a whole did not have higher utilization or payments than LEARN participants, whereas A4 Medicare participants with indicators of Alzheimer’s disease progression had greater health-care utilization and Medicare payments than those without such indicators. This reinforces the rationale for anchoring the present threshold exercise to delayed clinical progression rather than to biomarker status alone, while also underscoring that our analysis did not use observed claims or payment data.

Several limitations deserve emphasis. First, the translated benefits remain model-based counterfactual estimates rather than observed intervention effects in ADNI, A4, or LEARN (22,23). The analysis assumes that the association between cognitive slope and subsequent progression estimated in external cohorts can approximate the effect that would be observed if an intervention produced a comparable improvement in cognitive slope. This assumption may fail if cognitive slope is primarily a marker of underlying disease severity, comorbidity, selection, or other unmeasured causes of progression rather than a modifiable mediator on the pathway from intervention to delayed clinical conversion. Therefore, the estimates should be interpreted as clinical translations of a slope shift under explicit causal assumptions, not as proof that changing the slope by intervention would necessarily produce the same reduction in progression risk. A related issue is estimand alignment: the U.S. POINTER slope difference is a randomized between-group contrast, whereas the external-cohort slope-to-event associations were estimated conditional on baseline covariates including age, sex, education, and *APOE*-ε4. Without individual-level U.S. POINTER data, we could not fully align the marginal trial estimand with the adjusted external-cohort association structure. In addition, A4 was a randomized solanezumab trial rather than a purely observational natural-history cohort. Although the present study did not evaluate solanezumab efficacy and used A4 as an amyloid-elevated longitudinal progression dataset, treatment assignment and trial-specific procedures could still have influenced cognitive trajectories, follow-up patterns, or conversion ascertainment. We did not perform treatment-arm-specific or placebo-only A4 sensitivity analyses. This design feature should be considered when interpreting A4-based estimates. Finally, the cognitive composite used in U.S. POINTER and the cohort-specific mPACC or PACC-derived measures used in ADNI, A4, and LEARN were not identical instruments. Although all slopes were expressed on a standardized SD/year scale within each cohort, this procedure provides scale alignment rather than full psychometric equivalence. A +0.029 SD/year difference may therefore not have the same clinical meaning across composites if the tests differ in domain coverage, reliability, practice effects, ceiling effects, or sensitivity to preclinical AD. This limitation could bias the translated effects in either direction.

Second, the translated estimates were sensitive to analytic design, endpoint definition, and model structure. The A4 landmark analysis should be interpreted particularly cautiously because the eligible landmark risk set was small relative to the baseline cognitively normal sample, reflecting limited availability of assessments near the prespecified 24-month landmark. We used sustained conversion confirmed at the subsequent visit to reduce misclassification due to diagnostic fluctuation or reversion, but this definition may delay event timing or exclude transient conversions and thereby affect absolute risk, RD, NNT, and RMST estimates. We did not repeat the analyses using a single-visit conversion definition. The joint model used a slope-only association structure to align with the U.S. POINTER slope estimand, whereas alternative formulations based on current cognitive level, cumulative exposure, or combined level-and-slope associations could yield different translated effects. Future analyses with individual-level intervention data should compare slope-only, value-only, and value-plus-slope joint-model specifications. The ADNI reweighting sensitivity analysis aligned selected published U.S. POINTER baseline marginals only and could not recover the joint correlation structure among eligibility-defining baseline characteristics. We also did not model death as a competing risk; if mortality differed by cognitive trajectory or biomarker subgroup, treating death as non-informative censoring could affect absolute 5-year risk, RD, NNT, and RMST estimates. Finally, the economic scenarios were intentionally simplified and focused on incremental delivery costs rather than full societal or health-system costs. The WTP thresholds were illustrative assumptions and were not empirically derived disease-specific thresholds for preventing MCI/dementia conversion or gaining MCI-free years. These economic estimates are therefore best viewed as scenario-based summaries for interpreting the possible scale of value under explicit assumptions, not as definitive causal forecasts or within-trial cost-effectiveness evidence.

## 5. Conclusion

An incremental U.S. POINTER structured-versus-self-guided +0.029 SD/year difference in cognitive slope, when translated into ADNI, A4, and LEARN, corresponded to directionally consistent reductions in the predicted risk of progression from cognitively normal status to mild cognitive impairment or dementia. The magnitude of this translation depended on cohort event risk, biomarker/genotype enrichment, follow-up structure, and modeling framework. Joint longitudinal-survival models yielded larger translated effects than landmark Cox models, whereas absolute 5-year RMST gains were generally modest and were below 1 month in ADNI and LEARN.

These findings should not be interpreted as evidence against multidomain lifestyle intervention. Rather, they define the expected scale of clinical progression delay implied by the published U.S. POINTER cognitive-slope contrast under explicit model assumptions. The exploratory economic threshold analyses suggested that more favorable margins may arise in higher-risk ADNI subgroups, particularly those enriched for both *APOE*-ε4 and amyloid positivity, when incremental delivery costs are low and WTP assumptions are high. However, these economic findings are assumption-dependent threshold calculations and should not be interpreted as empirical cost-effectiveness evidence or as a basis for subgroup-targeted implementation. Future studies using individual-level intervention data, observed implementation costs, disease-specific health-economic outcomes, and target-population-specific assumptions are needed to determine the clinical and economic value of structured multidomain lifestyle interventions.

## Acknowledgements

Data collection and sharing for this project was funded by the Alzheimer’s Disease Neuroimaging Initiative (ADNI) (National Institutes of Health Grant U01 AG024904) and DOD ADNI (Department of Defense award number W81XWH-12-2-0012). ADNI is funded by the National Institute on Aging, the National Institute of Biomedical Imaging and Bioengineering, and through generous contributions from the following: AbbVie, Alzheimer’s Association; Alzheimer’s Drug Discovery Foundation; Araclon Biotech; BioClinica, Inc.; Biogen; Bristol-Myers Squibb Company; CereSpir, Inc.; Cogstate; Eisai Inc.; Elan Pharmaceuticals, Inc.; Eli Lilly and Company; EuroImmun; F. Hoffmann-La Roche Ltd and its affiliated company Genentech, Inc.; Fujirebio; GE Healthcare; IXICO Ltd.;Janssen Alzheimer Immunotherapy Research & Development, LLC.; Johnson & Johnson Pharmaceutical Research & Development LLC.; Lumosity; Lundbeck; Merck & Co., Inc.;Meso Scale Diagnostics, LLC.; NeuroRx Research; Neurotrack Technologies; Novartis Pharmaceuticals Corporation; Pfizer Inc.; Piramal Imaging; Servier; Takeda Pharmaceutical Company; and Transition Therapeutics. The Canadian Institutes of Health Research is providing funds to support ADNI clinical sites in Canada. Private sector contributions are facilitated by the Foundation for the National Institutes of Health (www.fnih.org). The grantee organization is the Northern California Institute for Research and Education, and the study is coordinated by the Alzheimer’s Therapeutic Research Institute at the University of Southern California. ADNI data are disseminated by the Laboratory for Neuro Imaging at the University of Southern California.

The authors acknowledge the A4 and LEARN Study Teams for their important contributions to the A4 and LEARN Studies. The A4 Study was funded by a public-private-philanthropic partnership, including the National Institutes of Health/National Institute on Aging (R01AG063689, U19AG010483, and U24AG057437), Eli Lilly and Company, the Alzheimer’s Association, the Accelerating Medicines Partnership through the Foundation for the National Institutes of Health, the GHR Foundation, the Davis Alzheimer Prevention Program, an anonymous foundation, and additional private donors to Brigham and Women’s Hospital, with in-kind support from Avid, Cogstate, Albert Einstein College of Medicine, and the Foundation for Neurologic Diseases. The companion observational Longitudinal Evaluation of Amyloid Risk and Neurodegeneration (LEARN) Study was funded by the Alzheimer’s Association and the GHR Foundation. The complete A4 and LEARN Study Team list is available through the ACTC A4 Study Team Lists page.

The authors’ affiliation, “*Dementia Inclusion and Therapeutics*,” is an endowed department funded by Effissimo Capital Management Pte Ltd.

AI-assisted tools were used for language editing; authors take full responsibility for the content.

## Consent Statement

Informed consent is not required for this type of study.

## Data Availability

The ADNI data are available through the ADNI repository and are shared via the LONI Image and Data Archive (IDA) (https://adni.loni.usc.edu/). The A4 Study data and the LEARN Study data are available from the A4/LEARN data sharing platform (https://www.a4studydata.org/).

## References

1. Baker LD, Espeland MA, Whitmer RA, et al. Structured vs Self-Guided Multidomain Lifestyle Interventions for Global Cognitive Function: The US POINTER Randomized Clinical Trial. JAMA 2025;334:681–691. 10.1001/jama.2025.12923

2. Ngandu T, Lehtisalo J, Solomon A, et al. A 2 year multidomain intervention of diet, exercise, cognitive training, and vascular risk monitoring versus control to prevent cognitive decline in at-risk elderly people (FINGER): a randomised controlled trial. Lancet 2015;385:2255–2263. 10.1016/S0140-6736(15)60461-5

3. Livingston G, Huntley J, Liu KY, et al. Dementia prevention, intervention, and care: 2024 report of the Lancet standing Commission. Lancet 2024;404:572–628. 10.1016/S0140-6736(24)01296-0

4. Baker LD, Snyder HM, Espeland MA, et al. Study design and methods: U.S. study to protect brain health through lifestyle intervention to reduce risk (U.S. POINTER). Alzheimers Dement 2024;20:769–782. 10.1002/alz.13365

5. Kivipelto M, Mangialasche F, Snyder HM, et al. World-Wide FINGERS Network: A global approach to risk reduction and prevention of dementia. Alzheimers Dement 2020;16:1078–1094. 10.1002/alz.12123

6. Uno H, Claggett B, Tian L, et al. Moving beyond the hazard ratio in quantifying the between-group difference in survival analysis. J Clin Oncol 2014;32:2380–2385. 10.1200/JCO.2014.55.2208

7. Donohue MC, Sperling RA, Salmon DP, et al. The preclinical Alzheimer cognitive composite: measuring amyloid-related decline. JAMA Neurol 2014;71:961–970. 10.1001/jamaneurol.2014.803

8. Hampton OL, Mukherjee S, Properzi MJ, et al. Harmonizing the preclinical Alzheimer cognitive composite for multicohort studies. Neuropsychology 2023;37:436–449. 10.1037/neu0000833

9. Yau WW, Kirn DR, Rabin JS, et al. Physical activity as a modifiable risk factor in preclinical Alzheimer’s disease. Nat Med 2025;31:4075–4083. 10.1038/s41591-025-03955-6

10. Sperling RA, Rentz DM, Johnson KA, et al. The A4 study: stopping AD before symptoms begin? Sci Transl Med 2014;6:228fs13. 10.1126/scitranslmed.3007941

11. Sperling RA, Donohue MC, Rissman RA, et al. Amyloid and Tau Prediction of Cognitive and Functional Decline in Unimpaired Older Individuals: Longitudinal Data from the A4 and LEARN Studies. J Prev Alzheimers Dis 2024;11:802–813. 10.14283/jpad.2024.122

12. Petersen RC, Aisen PS, Beckett LA, et al. Alzheimer’s Disease Neuroimaging Initiative (ADNI): clinical characterization. Neurology 2010;74:201–209. 10.1212/WNL.0b013e3181cb3e25

13. Neumann PJ, Kim DD. Cost-effectiveness Thresholds Used by Study Authors, 1990-2021. JAMA 2023;329:1312–1314. 10.1001/jama.2023.1792

14. Cameron D, Ubels J, Norström F. On what basis are medical cost-effectiveness thresholds set? Clashing opinions and an absence of data: a systematic review. Glob Health Action 2018;11:1447828. 10.1080/16549716.2018.1447828

15. Dafni U. Landmark analysis at the 25-year landmark point. Circ Cardiovasc Qual Outcomes 2011;4:363–371. 10.1161/CIRCOUTCOMES.110.957951

16. Henderson R, Diggle P, Dobson A. Joint modelling of longitudinal measurements and event time data. Biostatistics 2000;1:465–480. 10.1093/biostatistics/1.4.465

17. Wulfsohn MS, Tsiatis AA. A joint model for survival and longitudinal data measured with error. Biometrics 1997;53:330–339. 10.2307/2533118

18. Whitmer RA, Baker LD, Carrillo MC, et al. Baseline characteristics of the U.S. Study to Protect Brain Health Through Lifestyle Intervention to Reduce Risk (U.S. POINTER): Successful enrollment of a diverse clinical trial cohort at risk for cognitive decline. Alzheimers Dement 2025;21:e70351. 10.1002/alz.70351

19. Chen R, Chen G, Yu M. Entropy balancing for causal generalization with target sample summary information. Biometrics 2023;79:3179–3190. 10.1111/biom.13825

20. Boudewijns EA, Otten TM, Gobianidze M, Ramaekers BL, van Schayck OCP, Joore MA. Headroom Analysis for Early Economic Evaluation: A Systematic Review. Appl Health Econ Health Policy 2023;21:195–204. 10.1007/s40258-022-00774-5

21. Sanders GD, Neumann PJ, Basu A, et al. Recommendations for Conduct, Methodological Practices, and Reporting of Cost-effectiveness Analyses: Second Panel on Cost-Effectiveness in Health and Medicine. JAMA 2016;316:1093–1103. 10.1001/jama.2016.12195

22. Dahabreh IJ, Hernán MA. Extending inferences from a randomized trial to a target population. Eur J Epidemiol 2019;34:719–722. 10.1007/s10654-019-00533-2

23. Dahabreh IJ, Haneuse SJA, Robins JM, et al. Study Designs for Extending Causal Inferences From a Randomized Trial to a Target Population. Am J Epidemiol 2021;190:1632–1642. 10.1093/aje/kwaa270

24. Mormino EC, Papp KV. Amyloid Accumulation and Cognitive Decline in Clinically Normal Older Individuals: Implications for Aging and Early Alzheimer’s Disease. J Alzheimers Dis 2018;64:S633–S646. 10.3233/JAD-179928

25. Andrieu S, Guyonnet S, Coley N, et al. Effect of long-term omega 3 polyunsaturated fatty acid supplementation with or without multidomain intervention on cognitive function in elderly adults with memory complaints (MAPT): a randomised, placebo-controlled trial. Lancet Neurol 2017;16:377–389. 10.1016/S1474-4422(17)30040-6

26. Moll van Charante EP, Richard E, Eurelings LS, et al. Effectiveness of a 6-year multidomain vascular care intervention to prevent dementia (preDIVA): a cluster-randomised controlled trial. Lancet 2016;388:797–805. 10.1016/S0140-6736(16)30950-3

27. Brodaty H, Chau T, Heffernan M, et al. An online multidomain lifestyle intervention to prevent cognitive decline in at-risk older adults: a randomized controlled trial. Nat Med 2025;31:565–573. 10.1038/s41591-024-03351-6

28. Coley N, Hoevenaar-Blom MP, Shourick J, et al. Searching for responders to multidomain dementia prevention in late life: A pooled analysis of individual participant data from the MAPT and preDIVA trials. Alzheimers Dement 2025;21:e14472. 10.1002/alz.14472

29. Altman DG, Andersen PK. Calculating the number needed to treat for trials where the outcome is time to an event. BMJ 1999;319:1492–1495. 10.1136/bmj.319.7223.1492

30. Royston P, Parmar MK. Restricted mean survival time: an alternative to the hazard ratio for the design and analysis of randomized trials with a time-to-event outcome. BMC Med Res Methodol 2013;13:152. 10.1186/1471-2288-13-152

31. Wimo A, Handels R, Antikainen R, et al. Dementia prevention: The potential long-term cost-effectiveness of the FINGER prevention program. Alzheimers Dement 2023;19:999–1008. 10.1002/alz.12698

32. Welberry HJ, Ku LE, Shih ST, et al. The cost-effectiveness of an online intervention to prevent dementia: Results from the Maintain Your Brain (MYB) randomised controlled trial. J Prev Alzheimers Dis 2025;12:100151. 10.1016/j.tjpad.2025.100151

33. Takashi N, Ohtera S, Kuroda Y, Arai H, Sakurai T. Cost-effectiveness of multimodal intervention for the prevention of dementia in Japan. J Prev Alzheimers Dis 2026;13:100460. 10.1016/j.tjpad.2025.100460

34. Beyrer J, Sheff Z, Payakachat N, Chandler JM, Chen YF, Kubisiak J, Lee A, Holdridge KC, Yaari R, Aisen P, Rafii MS, Sperling RA; A4 and LEARN Study Teams. Increase in healthcare utilization and Medicare payment with progression of preclinical Alzheimer’s disease. J Prev Alzheimers Dis. 2026 Apr 1;13(6):100547. doi: 10.1016/j.tjpad.2026.100547. Epub ahead of print. PMID: 41926844; PMCID: PMC13084671.

35. Papp KV, Rentz DM, Orlovsky I, Sperling RA, Mormino EC. Optimizing the preclinical Alzheimer’s cognitive composite with semantic processing: The PACC5. Alzheimers Dement (N Y) 2017;3:668–677. 10.1016/j.trci.2017.10.004

36. Hansson O, Seibyl J, Stomrud E, et al. CSF biomarkers of Alzheimer’s disease concord with amyloid-β PET and predict clinical progression: A study of fully automated immunoassays in BioFINDER and ADNI cohorts. Alzheimers Dement 2018;14:1470–1481. 10.1016/j.jalz.2018.01.010

37. Royse SK, Minhas DS, Lopresti BJ, et al. Validation of amyloid PET positivity thresholds in centiloids: a multisite PET study approach. Alzheimers Res Ther 2021;13:99. 10.1186/s13195-021-00836-1

38. Sperling RA, Donohue MC, Raman R, et al. Association of Factors With Elevated Amyloid Burden in Clinically Normal Older Individuals. JAMA Neurol 2020;77:735–745. 10.1001/jamaneurol.2020.0387

