## Supplementary Materials for "Mapping U.S. POINTER Cognitive-Slope Gains Onto Predicted Clinical Progression: An External-Cohort Translation Analysis With Exploratory Economic Thresholds"

**Table S1. Proportional hazards assessment for Cox survival components used in the landmark and joint-model analyses**

| **Cohort** | **Framework** | **Events/N** | **Global Schoenfeld p** | **Minimum covariate p** | **Slope p** | **Age p** | **Sex p** | **Education p** | ***APOE*-**ε**4 p** |
| --- | --- | --- | --- | --- | --- | --- | --- | --- | --- |
| ADNI | Landmark model | 61/399 | 0.589 | 0.166 | 0.166 | 0.250 | 0.996 | 0.957 | 0.600 |
|  | Joint model | 86/486 | 0.421 | 0.132 | NA | 0.280 | 0.556 | 0.825 | 0.132 |
| A4 | Landmark model | 37/124 | 0.711 | 0.315 | 0.827 | 0.430 | 0.565 | 0.315 | 0.542 |
|  | Joint model | 410/1064 | 0.572 | 0.278 | NA | 0.350 | 0.995 | 0.465 | 0.278 |
| LEARN | Landmark model | 45/394 | 0.194 | 0.095 | 0.373 | 0.422 | 0.238 | 0.095 | 0.265 |
|  | Joint model | 87/505 | 0.603 | 0.127 | NA | 0.127 | 0.861 | 0.653 | 0.910 |

Scaled Schoenfeld residuals were used to assess proportional hazards assumptions in the Cox survival components. For the landmark analyses, diagnostics were applied to the fitted post-landmark Cox models. For the joint-model analyses, diagnostics were applied to the initial Cox survival components used to define the event process before fitting the Bayesian joint longitudinal-survival models. These diagnostics were interpreted descriptively and were not considered formal tests of the latent slope-association structure in the final joint models. Slope p-values are not applicable to the joint-model initial Cox components because participant-specific cognitive slope was not included as a fixed covariate in those initial Cox models.

Abbreviations: ADNI, Alzheimer's Disease Neuroimaging Initiative; A4, Anti-Amyloid Treatment in Asymptomatic Alzheimer's Disease; LEARN, Longitudinal Evaluation of Amyloid Risk and Neurodegeneration; *APOE*, apolipoprotein E; mPACC, modified Preclinical Alzheimer Cognitive Composite.

**Table S2. ADNI reweighting diagnostics for alignment to published U.S. POINTER parent-trial baseline marginals**

| Characteristic | Metric | ADNI observed | U.S. POINTER target | ADNI weighted | Absolute standardized difference before | Absolute standardized difference after |
| --- | --- | --- | --- | --- | --- | --- |
| Age | Mean (years) | 73.7 | 68.2 | 68.7 | 0.87 | 0.07 |
| Female sex | Proportion | 0.523 | 0.689 | 0.677 | 0.34 | 0.03 |
| Age >=70 years | Proportion | 0.752 | 0.430 | 0.447 | 0.69 | 0.03 |
| *APOE*-ε4 carrier | Proportion | 0.277 | 0.300 | 0.298 | 0.05 | <0.01 |
| Effective sample size | N | 512 | N/A | 254.6 | N/A | N/A |

Observed values are calculated in the complete-case ADNI baseline cognitively normal sample used for reweighting (n = 512). Weighted values are obtained after entropy balancing to selected published U.S. POINTER parent-trial baseline marginals.

The primary calibration targets were mean age = 68.2 years, female proportion = 0.689, age >=70 years proportion = 0.43, and *APOE*-ε4 carrier proportion = 0.30.

Absolute standardized differences were calculated using the observed ADNI standard deviation for the continuous age variable and the pooled binomial standard deviation for binary variables.

Effective sample size was calculated as (sum w_i)^2 / sum w_i^2.

These weights align selected published U.S. POINTER parent-trial marginals only and do not reconstruct individual-level U.S. POINTER participants or the joint correlation structure among baseline characteristics.

Amyloid positivity was not included as a primary calibration target because the publicly available 29.2% value corresponded to the imaging ancillary cohort rather than the full randomized parent trial.

Abbreviations: ADNI, Alzheimer’s Disease Neuroimaging Initiative; *APOE*, apolipoprotein E.

**Table S3. Economic translation formulas and scenario definitions**

| **Quantity** | **Definition / Formula** | **Unit** | **Interpretation** |
| --- | --- | --- | --- |
| 5-year risk difference (RD) | RD = Risk_observed - Risk_shifted | percentage points | Positive values indicate lower predicted 5-year conversion risk under the +0.029 SD/year shifted scenario. |
| 5-year number needed to treat (NNT) | NNT = 1 / RD | persons treated for 5 years | Interpreted only when RD is positive and sufficiently far from zero. |
| 5-year RMST difference | RMST_diff = RMST_shifted - RMST_observed | months | Positive values indicate longer predicted time free from sustained conversion under the shifted scenario. |
| RMST difference in years | RMST_diff,year = RMST_diff,month / 12 | years | Used for MCI-free-year-based economic translation. |
| Maximum allowable cost, case-prevention basis | MaxCost_case = WTP_case x RD | USD/person over 2 years | Equivalent to WTP_case / NNT when RD is expressed as an absolute probability difference. |
| Maximum allowable cost, MCI-free-year basis | MaxCost_year = WTP_year x RMST_diff,year | USD/person over 2 years | Uses translated RMST gain as the measure of MCI-free time gained. |
| Net monetary benefit, case-prevention basis | NMB_case = MaxCost_case - Cost_program | USD/person | Positive values indicate that translated case-prevention benefit exceeds the assumed incremental program cost. |
| Net monetary benefit, MCI-free-year basis | NMB_year = MaxCost_year - Cost_program | USD/person | Positive values indicate that translated MCI-free-year benefit exceeds the assumed incremental program cost. |
| Break-even incremental cost | Break-even Cost = MaxCost | USD/person over 2 years | Incremental cost at which net monetary benefit equals zero. |

All economic quantities were derived from translated clinical-effect estimates rather than from observed within-trial cost, utility, or resource-use data. The economic analysis was therefore scenario-based and should not be interpreted as a within-trial cost-effectiveness analysis.

The primary incremental program-cost scenario was 400 USD per participant over 2 years. Sensitivity analyses used 600 and 800 USD per participant over 2 years.

The primary case-based willingness-to-pay threshold was 100,000 USD per case prevented, with 150,000 USD per case prevented in sensitivity analyses. The MCI-free-year-based summary used 100,000 USD per MCI-free year.

For the case-prevention summary, RD should be expressed as an absolute probability difference per participant over 5 years when used in the formula MaxCost_case = WTP_case x RD. When RD is reported in percentage points in the main tables, conversion to probability units is required before applying this formula.

NNT becomes unstable when RD is close to zero; accordingly, case-based economic summaries from landmark analyses with near-null risk differences should be interpreted cautiously.

A positive NMB indicates that the translated clinical benefit would justify the assumed incremental intervention cost under the selected WTP threshold; a negative NMB indicates that it would not.

ADNI subgroup economic analyses were restricted to the Overall, *APOE*-ε4+, amyloid-positive, and *APOE*-ε4+ plus amyloid-positive groups because A4 and LEARN were structurally defined by amyloid-screening eligibility and therefore were not suitable for balanced subgroup economic comparison.

Abbreviations: RD, risk difference; NNT, number needed to treat; RMST, restricted mean survival time; WTP, willingness-to-pay; NMB, net monetary benefit.

**Table S4. Distribution of landmark-window selection modes across cohorts**

| **Cohort** | **Closest visit on/after Month 24 within Month 24-36, n (%)** | **Nearest prior visit within Month 12 to <24, n (%)** | **Total landmark analytic sample, N** |
| --- | --- | --- | --- |
| **ADNI** | 389 (97.5) | 10 (2.5) | 399 |
| **A4** | 1 (0.8) | 123 (99.2) | 124 |
| **LEARN** | 361 (91.6) | 33 (8.4) | 394 |

Percentages are calculated among participants included in the final landmark analytic sample within each cohort and may not sum to 100 because of rounding.

Abbreviations: ADNI, Alzheimer's Disease Neuroimaging Initiative; A4, Anti-Amyloid Treatment in Asymptomatic Alzheimer's Disease; LEARN, Longitudinal Evaluation of Amyloid Risk and Neurodegeneration.

**Table S5. Joint-model time-function choices and convergence diagnostics**

| **Cohort** | **Events/N** | **Median visits** | **Time function** | **Max R-hat** | **Median ESS** |
| --- | --- | --- | --- | --- | --- |
| ADNI | 86/486 | 5 | Linear time | 1.030 | 500 |
| A4 | 410/1064 | 4 | Linear time | 1.002 | 1129 |
| LEARN | 87/505 | 6 | Linear time | 1.006 | 511 |

R-hat and effective sample size (ESS) were summarized across monitored posterior parameters extracted from the final JMbayes2 joint-model fits. All models used a slope-only association structure.

Abbreviations: ADNI, Alzheimer's Disease Neuroimaging Initiative; A4, Anti-Amyloid Treatment in Asymptomatic Alzheimer's Disease; LEARN, Longitudinal Evaluation of Amyloid Risk and Neurodegeneration; ESS, effective sample size.

**Table S6. Sensitivity analysis propagating uncertainty in the U.S. POINTER reference slope increment**

**A. Cohort-level translation**

| **Cohort** | **Model** | **HR (95% interval)** | **RD (95% interval), pp** | **NNT (95% interval)** | **RMST (95% interval), month** | **Pr (RD > 0)** |
| --- | --- | --- | --- | --- | --- | --- |
| ADNI | Landmark model | 0.974 (0.938-0.996) | 0.29 (0.04-0.70) | 345.9 (142.7-2035.8) | 0.057 (0.007-0.143) | 0.992 |
|  | Joint model | 0.835 (0.707-0.953) | 1.23 (0.33-2.33) | 81.0 (42.8-284.2) | 0.291 (0.078-0.551) | 0.997 |
| A4 | Landmark model | 0.998 (0.988-1.006) | 0.05 (-0.15-0.31) | Unstable | 0.011 (-0.038-0.071) | 0.723 |
|  | Joint model | 0.917 (0.860-0.977) | 3.02 (0.84-5.30) | 33.0 (18.9-112.4) | 1.236 (0.342-2.169) | 0.997 |
| LEARN | Landmark model | 0.996 (0.987-1.002) | 0.07 (-0.04-0.25) | Unstable | 0.018 (-0.012-0.064) | 0.891 |
|  | Joint model | 0.840 (0.688-0.961) | 2.15 (0.53-4.50) | 46.4 (22.2-176.5) | 0.591 (0.145-1.235) | 0.996 |

**B. ADNI subgroup translation**

| **Group** | **Model** | **Events/N** | **HR (95% interval)** | **RD (95% interval), pp** | **NNT (95% interval)** | **RMST (95% interval), month** | **Pr (RD > 0)** |
| --- | --- | --- | --- | --- | --- | --- | --- |
| Overall | Landmark model | 61/399 | 0.974 (0.938-0.996) | 0.29 (0.04-0.69) | 343.0 (144.8-1880.4) | 0.058 (0.007-0.142) | 0.994 |
|  | Joint model | 86/486 | 0.834 (0.707-0.954) | 1.23 (0.33-2.33) | 81.1 (42.8-284.8) | 0.291 (0.078-0.551) | 0.997 |
| *APOE*-ε4+ | Landmark model | 23/107 | 0.979 (0.927-1.014) | 0.38 (-0.20-1.29) | Unstable | 0.049 (-0.041-0.185) | 0.906 |
|  | Joint model | 33/133 | 0.841 (0.663-0.972) | 1.84 (0.36-4.21) | 54.2 (23.8-253.2) | 0.462 (0.090-1.047) | 0.996 |
| Amyloid+ | Landmark model | 38/128 | 0.965 (0.917-0.996) | 0.54 (0.08-1.30) | 183.8 (76.9-1026.2) | 0.101 (0.004-0.262) | 0.992 |
|  | Joint model | 48/161 | 0.822 (0.672-0.952) | 1.64 (0.43-3.26) | 60.7 (30.6-218.0) | 0.382 (0.100-0.761) | 0.997 |
| *APOE*-ε4+ & Amyloid+ | Landmark model | 17/46 | 0.979 (0.901-1.043) | 0.37 (-0.66-1.78) | Unstable | 0.049 (-0.124-0.274) | 0.781 |
|  | Joint model | 22/61 | 0.790 (0.550-0.982) | 2.48 (0.37-5.92) | 40.2 (16.9-221.5) | 0.677 (0.105-1.611) | 0.993 |

NNT was labeled unstable when a non-negligible proportion of Monte Carlo draws yielded a non-positive risk difference. The U.S. POINTER reference slope increment was treated as Δ_POINTER ~ Normal(0.029, SE_POINTER²), with SE_POINTER derived from the published 95% confidence interval for the U.S. POINTER between-group slope difference.

Abbreviations: ADNI, Alzheimer's Disease Neuroimaging Initiative; A4, Anti-Amyloid Treatment in Asymptomatic Alzheimer's Disease; LEARN, Longitudinal Evaluation of Amyloid Risk and Neurodegeneration; HR, hazard ratio; RD, risk difference; NNT, number needed to treat; RMST, restricted mean survival time.

**Supplementary Methods**

**#1. Construction and scale alignment of cohort-specific cognitive composites**

The longitudinal cognitive outcome was a cohort-specific mPACC or PACC-derived composite. The original PACC was developed for preclinical Alzheimer’s disease trials to capture subtle early cognitive decline by combining episodic memory, timed executive function or processing speed, and global cognitive performance (7). Because the available cognitive batteries differed across ADNI, A4, and LEARN, we did not impose an identical item-level composite across cohorts. Instead, we constructed or retained the best available cohort-specific PACC-compatible longitudinal cognitive measure and then applied cohort-specific standardization for the translation analyses. This approach is consistent with prior work showing that PACC-type composites can be adapted across cohorts, while also emphasizing that harmonization does not necessarily imply full psychometric equivalence (8,35).

In ADNI, mPACC was reconstructed from item-level cognitive tests. For ADNI1 participants, the composite included RAVLT immediate recall, Logical Memory delayed recall (LDELTOTAL), MMSE, and Digit Symbol score. For non-ADNI1 participants, Trail Making Test Part B replaced Digit Symbol. Trail Making Test Part B was winsorized at the 99th percentile and sign-reversed so that higher values indicated better cognition. For ADNI, each component was standardized using the baseline cognitively normal cohort mean and standard deviation for that component. The standardized components were then averaged to create a visit-level composite score. The composite was subsequently re-standardized using the baseline composite mean and standard deviation. Finally, each participant’s baseline standardized composite value was subtracted from all subsequent values, so that the ADNI mPACC represented within-person change from baseline, with higher values indicating better cognitive performance or less decline. Visits with insufficient component data to calculate the prespecified composite were treated as missing; no item-level imputation was performed.

In A4 and LEARN, we used the ACTC-derived PACC variable from the PACC.csv dataset. This variable was already defined as change from baseline PACC.raw, and therefore was used as the mPACC-equivalent longitudinal outcome without reconstructing item-level components. A4 and LEARN were designed as complementary cohorts of cognitively unimpaired older adults with different amyloid-risk profiles, and their longitudinal cognitive outcomes were therefore suitable for examining how a common standardized cognitive-slope shift translated across distinct biomarker-risk settings (10,11). Screening Clinical Dementia Rating information was carried forward to the baseline PACC visit when needed to align cognitive and clinical status at baseline, and analytic observations were restricted to visits with both PACC and clinical diagnostic information available.

Across cohorts, the resulting cognitive outcome should be interpreted as a cohort-specific PACC-compatible longitudinal measure rather than a psychometrically identical test battery. The cohort-specific standardization aligned the scale of cognitive change for the purpose of translating a standardized slope increment into clinical progression metrics, but it did not establish item-level, latent-trait, or intervention-response equivalence across U.S. POINTER, ADNI, A4, and LEARN (7,8). Accordingly, the transported quantity in this study was a standardized cognitive-slope increment expressed in SD/year, not an assumption that the same cognitive instrument was administered or that the same cognitive domains were measured with identical sensitivity in all cohorts.

**#2. Biomarker definitions and amyloid classification**

Amyloid status was defined according to the original cohort-specific biomarker framework and was used for descriptive summaries and ADNI subgroup definitions. It was not included as a modeling variable in the primary cross-cohort translation analyses.

In ADNI, baseline amyloid positivity was determined using the biomarker assessment closest to Month 0 within a ±3-month window. Participants were classified as amyloid-positive if either CSF or amyloid PET was positive. In the ADNI-based harmonized analytic dataset used for this study, CSF amyloid positivity was operationally defined as CSF Aβ42 <880, consistent with prior ADNI analyses comparing CSF Aβ42 with florbetapir PET (36). PET positivity was based on available tracer-specific ADNI PET-core thresholds, including florbetapir (AV45; SUVR >1.11) and florbetaben (FBB; SUVR >1.08) (37). Because ADNI biomarker data were obtained from multiple modalities and data versions, this definition should be interpreted as an operational ADNI amyloid definition rather than as a single uniform biomarker assay.

In A4 and LEARN, amyloid status was based on the screening florbetapir PET classification used for trial eligibility (38). A4 enrolled cognitively normal individuals with elevated amyloid on screening PET, whereas LEARN enrolled otherwise similar individuals without elevated amyloid. For the A4/LEARN PET derivation in the harmonized dataset, positivity was coded from composite SUVR and visual read: SUVR ≥1.15 was classified as positive, SUVR <1.10 was classified as negative, and values from 1.10 to <1.15 were resolved by visual read.

**#3. Calculation of translated 5-year risk, risk difference, number needed to treat, and RMST**

For both analytic frameworks, the translated clinical effect of interest was defined by comparing two counterfactual scenarios. In the observed-slope scenario, each participant retained the cognitive trajectory implied by the fitted model. In the shifted-slope scenario, the participant-specific cognitive slope was improved by +0.029 SD/year, corresponding to the U.S. POINTER structured-versus-self-guided reference increment.

The primary absolute time horizon was 5 years after risk-set entry, corresponding to 60 months. For each scenario, we estimated the marginal cumulative risk of sustained conversion from cognitively normal status to MCI or dementia by first obtaining model-based survival predictions and then averaging these predictions over the analytic sample.

The 5-year risk under the observed-slope scenario was defined as: *Risk_observed(5 years) = 1 − mean_i{S_observed,i(5 years)}*. The 5-year risk under the shifted-slope scenario was defined as: *Risk_shifted(5 years) = 1 − mean_i{S_shifted,i(5 years)}*. The translated 5-year risk difference was defined as: *RD = Risk_observed(5 years) − Risk_shifted(5 years)*. Positive RD values indicate lower predicted 5-year conversion risk under the +0.029 SD/year shifted-slope scenario.

The number needed to treat was defined as: *NNT = 1 / RD*, when RD was expressed as an absolute probability difference and was positive. When RD was non-positive, close to zero, or when the uncertainty interval included zero, NNT was treated as unstable and interpreted cautiously.

Restricted mean survival time over 5 years was defined as the area under the corresponding survival curve from 0 to 60 months: *RMST = ∫_0^60 S(t) dt*. The translated RMST difference was defined as: *RMST_diff = RMST_shifted − RMST_observed*. Positive RMST_diff values indicate a longer predicted duration free from sustained conversion under the shifted-slope scenario. RMST differences were reported in months. For economic translation on an MCI-free-year basis, RMST_diff in months was divided by 12 to express the gain in years.

**#4. Landmark Cox analysis**

For the landmark analysis, eligibility was defined at a Month 24 target landmark. Participants entered the landmark risk set only if they remained cognitively normal through that landmark. The landmark visit was defined as the assessment closest to Month 24 within a prespecified window. We first searched for a visit between Month 24 and Month 36 and selected the one closest to Month 24. If no such visit was available, we used the nearest prior visit between Month 12 and less than Month 24.

Within each cohort, the longitudinal cognitive outcome was standardized to a z score before slope estimation. For participant i at visit j, the early cognitive trajectory was modeled as: *mPACC_z,ij = β0 + b0i + (β1 + b1i)t_ij + ε_ij*, where t_ij denotes time in years from baseline, β0 and β1 are fixed effects, b0i is the participant-specific random intercept, b1i is the participant-specific random slope, and ε_ij is residual error. In the prespecified analysis pipeline, the random intercept and slope were specified with zero covariance. If this model was unstable or singular, participant-specific ordinary least-squares slopes were used as a fallback.

The estimated annual slope for participant i was then entered into a Cox proportional hazards model for time from landmark entry to sustained conversion: *h_i(t) = h_0(t) exp(θ1 slope_i + θ2 age_i + θ3 sex_i + θ4 education_i + θ5 APOE-ε4_i)*. The translated hazard ratio for a +0.029 SD/year improvement in cognitive slope was calculated as: *HR_Δ = exp(θ1 × 0.029)*. The observed-slope and shifted-slope scenarios were then evaluated using the fitted Cox model. In the shifted-slope scenario, each participant’s estimated annual cognitive slope was increased by +0.029 SD/year. Marginal survival curves were obtained under both scenarios, averaged across participants, and used to derive 5-year risk, RD, NNT, and RMST difference as described above.

Uncertainty for landmark-derived translated estimates was quantified using participant-level bootstrap resampling. Resampling was performed at the participant level, with replacement, separately within each cohort and, for subgroup analyses, within each ADNI subgroup. Within each bootstrap sample, the full landmark pipeline was rerun: early cognitive slopes were re-estimated, the post-landmark risk set was reconstructed using the prespecified landmark eligibility rules, the Cox model was refit, and the observed-slope and shifted-slope scenarios were recomputed. Percentile-based 95% confidence intervals were obtained from 500 bootstrap resamples.

The proportional hazards assumption for the landmark Cox models was assessed using scaled Schoenfeld residuals. Results are summarized in Table S1.

**#5. Joint longitudinal–survival model**

For the joint longitudinal–survival analysis, longitudinal mPACC observations from baseline through Month 84 were modeled jointly with time to sustained conversion. Time since baseline was expressed in years and centered at the cohort-specific mean to improve numerical stability. The longitudinal submodel was: *mPACC_z,ij = m_i(t_ij) + ε_ij*, with *m_i(t) = β0 + β1 t + β2 age_i + β3 sex_i + β4 education_i + β5 APOE-ε4_i + b0i + b1i t*, where m_i(t) denotes the latent participant-specific cognitive trajectory, b0i is the participant-specific random intercept, and b1i is the participant-specific random slope. The default time effect was linear. If needed for stable fitting, natural spline terms with 2 or 3 degrees of freedom were allowed within the prespecified analysis pipeline.

The survival submodel was: *h_i(t) = h_0(t) exp(γ^T Z_i + α m_i'(t))*, where Z_i contains baseline covariates, including age, sex, education, and *APOE*-ε4, and m_i'(t) denotes the instantaneous slope of the latent cognitive trajectory. The parameter α represents the association between the latent instantaneous cognitive slope and the hazard of sustained conversion. We used a slope-only association structure because the scientific question of interest was how a difference in the rate of cognitive change would translate into a difference in progression risk.

The model was estimated in a Bayesian framework using Markov chain Monte Carlo sampling as implemented in *JMbayes2*. In the final analysis, four chains were run with 20,000 iterations per chain, a burn-in of 5,000 iterations, and thinning of 2. If the maximum R-hat exceeded 1.10, a longer refit with 30,000 iterations per chain was prespecified.

For translation of the U.S. POINTER benchmark, posterior draws of the slope-association parameter α were transformed into posterior draws of the translated hazard ratio: *HR_Δ = exp(α × 0.029)*. For each posterior draw, model-based dynamic predictions were used to compute survival probabilities under the observed-slope and +0.029 SD/year shifted-slope scenarios. These posterior predictive survival trajectories were then used to calculate 5-year risk, RD, NNT, and RMST difference. Point estimates and 95% credible intervals were obtained directly from the posterior distribution of each translated quantity.

Convergence diagnostics, including maximum R-hat and effective sample size summaries, are reported in Table S5. The proportional hazards assumption for the initial Cox survival components used before fitting the Bayesian joint models was assessed descriptively using Schoenfeld residuals and is summarized in Table S1. These diagnostics were not considered formal tests of the latent slope-association structure in the final joint models.

**#6. Propagation of uncertainty in the U.S. POINTER reference slope increment**

The primary analyses treated the published U.S. POINTER structured-versus-self-guided slope difference of +0.029 SD/year as a fixed reference increment. Because this value is itself an estimate, we performed an approximate Monte Carlo sensitivity analysis that propagated uncertainty in the U.S. POINTER reference effect.

The U.S. POINTER reference increment was modeled as: *Δ_POINTER ~ Normal(0.029, SE_POINTER²)*, where SE_POINTER was derived from the published 95% confidence interval for the U.S. POINTER between-group slope difference. If the published 95% confidence interval is denoted by [L, U], then: *SE_POINTER = (U − L) / (2 × 1.96)*. Using the published confidence interval of 0.008 to 0.050 SD/year: *SE_POINTER = (0.050 − 0.008) / (2 × 1.96)* ≈ 0.0107. Because individual-level U.S. POINTER data were not available, this sensitivity analysis did not refit the individual-level external-cohort models for each draw of Δ_POINTER. Instead, it used the primary estimates and intervals from Tables 2 and 3 to approximate external-cohort slope-to-event uncertainty.

For HRs, we first derived the implied log-HR per 1 SD/year slope increment from the primary translated HR: *β_slope = log(HR_primary) / 0.029*. For each Monte Carlo draw k, we calculated: *HR_k = exp(β_slope,k × Δ_POINTER,k)*, where β_slope,k incorporated uncertainty approximated from the reported primary interval for the translated HR.

For RD and RMST, we used a local linear approximation around Δ_POINTER = 0.029. Specifically, if Q_primary denotes the primary translated RD or RMST difference at Δ_POINTER = 0.029, then for each Monte Carlo draw k: *Q_k ≈ Q_primary,k × (Δ_POINTER,k / 0.029)*, where Q_primary,k incorporated uncertainty approximated from the reported primary interval for that quantity. This approximation assumes local linearity of RD and RMST with respect to the small slope shift around the primary U.S. POINTER benchmark.

For each cohort and ADNI subgroup, Monte Carlo distributions were summarized using point estimates and 95% intervals. NNT was calculated only when the corresponding RD draw was positive. NNT was labeled unstable when a non-negligible proportion of Monte Carlo draws yielded a non-positive RD. We also reported Pr(RD > 0), the proportion of Monte Carlo draws in which the translated RD was positive. Results are shown in Table S6.

**#7. ADNI entropy-balancing sensitivity analysis**

Because the primary cross-cohort analyses asked how a U.S. POINTER-sized standardized slope improvement would translate within the observed baseline distributions of ADNI, A4, and LEARN, rather than by reconstructing POINTER-like individuals inside those cohorts, we performed an ADNI-only sensitivity analysis based on marginal realignment to the publicly reported U.S. POINTER parent-trial baseline characteristics.

We used entropy-balancing weights to align the ADNI baseline cognitively normal sample to selected published U.S. POINTER parent-trial marginals. The calibration targets were mean age = 68.2 years, female proportion = 0.689, age 70 years or older proportion = 0.43, and *APOE*-ε4 carrier proportion = 0.30. Amyloid positivity was not used as a calibration target because the publicly available 29.2% value corresponded to the imaging ancillary cohort rather than the full randomized parent trial. The entropy-balancing procedure was designed to preserve as much of the original ADNI sample as possible while forcing agreement with the selected target moments.

For landmark analyses, the calibration weights were applied directly in the Cox-model translation step. For the joint-model sensitivity analysis, because the JMbayes2 implementation used here did not directly support participant-level calibration weights in the full longitudinal-survival fitting step, we generated a weighted pseudo-population by stochastic replication proportional to the entropy-balancing weights and then refit the joint model in that pseudo-population. This approach should be interpreted as a sensitivity analysis based on selected baseline marginals only and not as a reconstruction of the individual-level U.S. POINTER participant distribution.

To summarize the degree of realignment, we reported the observed ADNI baseline moments, the published U.S. POINTER targets, the weighted ADNI moments, absolute standardized differences before and after weighting, and the effective sample size of the reweighted cohort. The effective sample size was calculated as: *ESS = (Σw_i)^2 / Σw_i^2*. Reweighting diagnostics are shown in Table S2.

**#8. Economic translation formulas and scenario assumptions**

The economic component was designed as an exploratory threshold exercise and not as a within-trial cost-effectiveness analysis. We did not estimate quality-adjusted life-years or use observed cost, utility, resource-use, or implementation-cost data from U.S. POINTER. Instead, we mapped the translated clinical effect measures obtained in ADNI subgroups to maximum allowable cost and net monetary benefit under prespecified incremental-cost and willingness-to-pay assumptions.

Economic analyses were restricted to ADNI because A4 and LEARN were structurally defined by amyloid-screening eligibility and therefore did not provide balanced subgroup contrasts for biomarker-based economic comparison. Four ADNI groups were evaluated: Overall, *APOE*-ε4 carriers, amyloid-positive participants, and *APOE*-ε4 plus amyloid-positive participants.

The primary incremental program-cost scenario was set to $400 per participant over 2 years, reflecting an illustrative low incremental delivery-cost margin for the additional intensity of the structured versus self-guided U.S. POINTER program. This difference was operationalized as 32 additional group meetings over 2 years, 2 additional coaching contacts, and a modest allowance for added administration and staffing. Sensitivity analyses used incremental costs of $600 and $800 over 2 years. These values were not intended to represent the full cost of implementing a multidomain lifestyle intervention.

The primary case-based willingness-to-pay threshold was $100,000 per prevented progression event, with $150,000 per prevented progression event examined in sensitivity analyses. We also summarized value on an MCI-free-year basis using $100,000 per MCI-free year. These WTP thresholds were used only as illustrative scale-setting assumptions. Because conventional cost-effectiveness thresholds are typically expressed per quality-adjusted life-year, they are not dimensionally equivalent to WTP per prevented MCI/dementia progression event or per MCI-free year.

For the case-prevention summary, the maximum allowable cost per participant was calculated as: *MaxCost_case = WTP_case × RD*, where RD was expressed as an absolute probability difference over 5 years. Equivalently, when RD was positive and sufficiently far from zero: *MaxCost_case = WTP_case / NNT*. For the MCI-free-year summary, the RMST gain was first converted from months to years: *RMST_diff,year = RMST_diff,month / 12*. The maximum allowable cost per participant on the MCI-free-year basis was then calculated as: *MaxCost_year = WTP_year × RMST_diff,year*. Net monetary benefit was defined as maximum allowable cost minus the assumed incremental program cost: *NMB_case = MaxCost_case − Cost_program, NMB_year = MaxCost_year − Cost_program*. Under this framework, positive net monetary benefit indicates that the translated clinical gain would justify the assumed incremental program cost under the selected WTP threshold, whereas negative net monetary benefit indicates that it would not. These quantities should be interpreted as scenario-based economic summaries derived from translated NNT- and RMST-based benefit estimates rather than as empirical estimates of observed health-system or societal cost savings. Economic formulas and scenario definitions are summarized in Table S3.

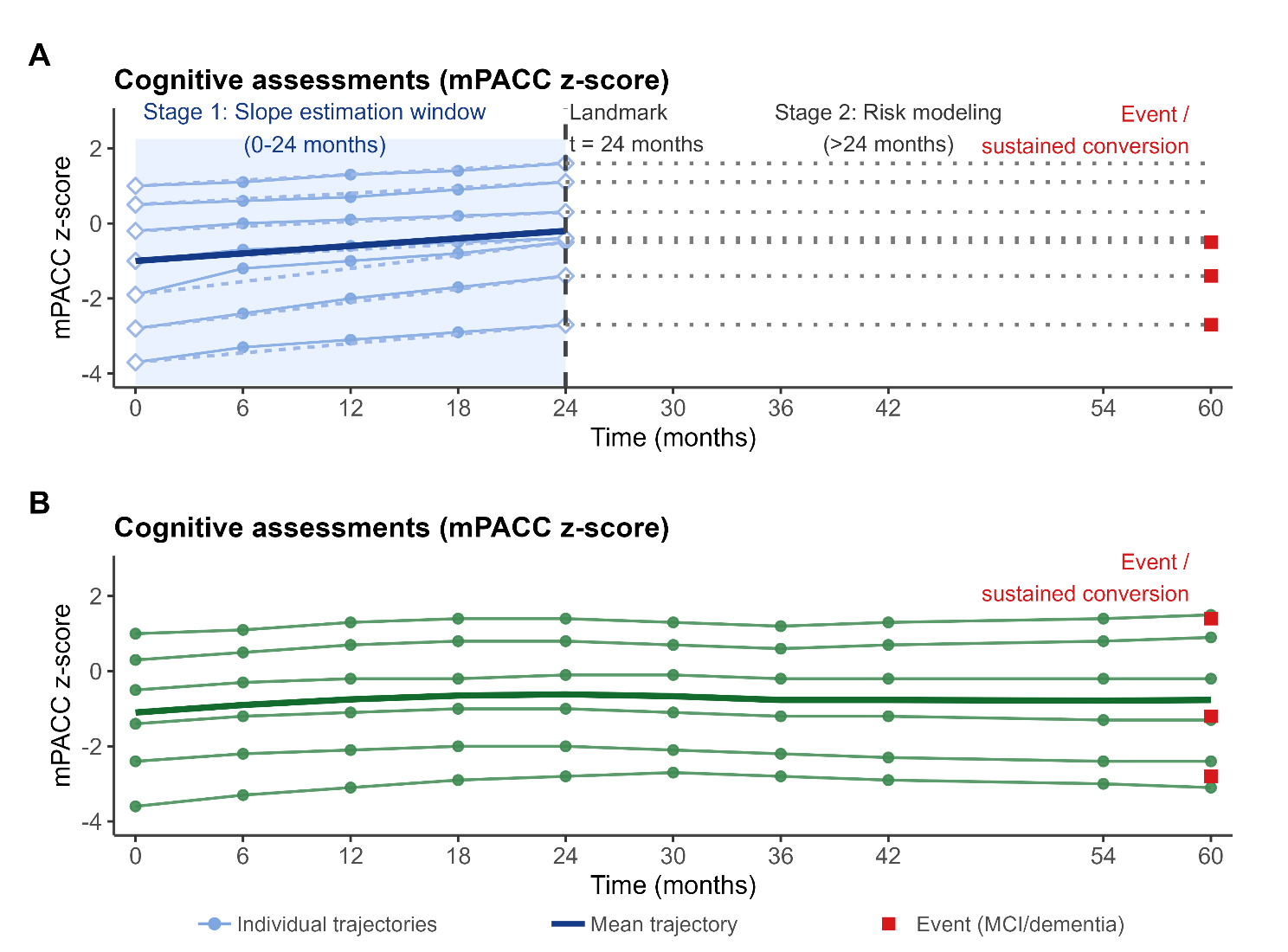
**Figure S1. Schematic overview of the two modeling approaches used to translate cognitive slope improvement into delayed clinical progression**

The figure illustrates how longitudinal cognitive assessments were linked to subsequent clinical progression in the two analytical frameworks. Thin lines represent individual mPACC trajectories, the thick line represents the mean trajectory, and red squares indicate sustained conversion to MCI or dementia. The figure is schematic and does not represent observed data.

**A. Landmark Cox model.** Cognitive change is first summarized during an initial slope-estimation window from baseline to 24 months. At the 24-month landmark, participants who remain cognitively normal enter the post-landmark risk set. The estimated individual cognitive slope is then used as a fixed predictor in a Cox model for subsequent sustained conversion to MCI or dementia.

**B. Joint longitudinal–survival model.** Repeated cognitive assessments across follow-up are modeled jointly with the time-to-conversion process. This approach uses the available longitudinal trajectory rather than reducing cognition to a single pre-landmark slope estimate, and links the latent cognitive trajectory to the risk of sustained conversion.

**Abbreviations:** CN, cognitively normal; MCI, mild cognitive impairment; mPACC, modified Preclinical Alzheimer Cognitive Composite; τ, landmark time.

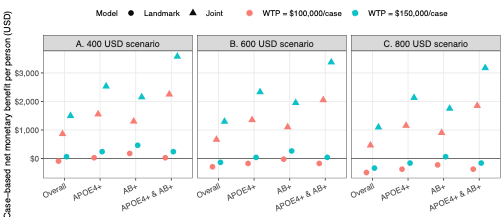
**Figure S2. Economic sensitivity analyses for ADNI subgroup translated effects**

This supplementary figure evaluates the robustness of the case-based economic interpretation of the ADNI subgroup results across alternative intervention cost and willingness-to-pay assumptions. The clinical input is the same as in Figure 1: a counterfactual +0.029 SD/year improvement in mPACC slope applied to ADNI subgroups, with translated effects estimated using landmark Cox and joint longitudinal–survival models. Panels show fixed incremental intervention cost scenarios over 2 years: A, $400 per person; B, $600 per person; and C, $800 per person. The y-axis shows case-based net monetary benefit (NMB) per person in USD, calculated as: *NMB = (WTP per prevented case × 5-year RD) − incremental cost, equivalently WTP/NNT − incremental cost when RD is positive*. Two willingness-to-pay thresholds are shown: $100,000 per prevented case and $150,000 per prevented case. Point shape indicates the statistical model, with circles for the landmark model and triangles for the joint model. Color indicates the willingness-to-pay threshold. The horizontal line at zero represents cost neutrality. Values above zero indicate positive net monetary benefit under the corresponding cost and willingness-to-pay scenario, whereas values below zero indicate that the assumed cost exceeds the monetized case-prevention benefit. Across scenarios, higher WTP thresholds and lower assumed costs yield higher NMB, and joint-model estimates generally produce larger NMB than landmark estimates.
